# Plasticity of the face-hand sensorimotor circuits after a traumatic brachial plexus injury

**DOI:** 10.1101/2022.10.13.22281048

**Authors:** Fernanda de Figueiredo Torres, Bia Lima Ramalho, Marcelle Ribeiro Rodrigues, Ana Carolina Schmaedeke, Victor Hugo Moraes, Karen T. Reilly, Raquel de Paula Carvalho, Claudia D. Vargas

## Abstract

**Background:** Traumatic brachial plexus injury (TBPI) is a potentially debilitating event, that usually affects young men following car or motorbike accidents. TBPI interferes with hand sensorimotor function, is associated with chronic pain, and causes cortical reorganization. Interactions between the somatosensory and motor cortices are of fundamental importance for motor control. The hands and face stand out as regions of high functionality with a privileged interaction existing between them, as reflected by the proximity and extension of their representations. Face-hand sensorimotor interactions have been demonstrated in healthy subjects.

**Objective:** The aim of this study was to investigate changes in the sensorimotor interaction in the hand and between the face and the hand in TBPI patients in order to better understand the plasticity of face-hand sensorimotor circuits following TBPI.

**Method:** The experimental design consisted of activating the representation of a hand muscle using transcranial magnetic stimulation (TMS) preceded by an electrical stimulation (ES) applied to the hand or face, which allows the investigation of the cortical reorganization resulting from TBPI. In the paradigm called afferent inhibition (AI), the motor evoked potential (MEP) in a target muscle is significantly reduced by a previous peripheral ES. AI can be evoked in short-latency (SAI) or long-latency (LAI) interstimulus intervals. Nine TBPI patients participated: five had partial sensorimotor function in their hands and were evaluated on the injured side (TBPI-I group) and four had complete loss of sensorimotor function in their hands and were evaluated on the uninjured side (TBPI-UI group). A control group (CG) included 18 healthy adults. A detailed clinical evaluation complemented the analysis.

**Results:** The results showed preserved hand sensorimotor integration for TBPI patients at SAI intervals, but not at LAI intervals. For the face-to-hand sensorimotor integration, the results showed no inhibition at SAI intervals for the TBPI patients. For LAI intervals, a facilitation effect was observed for the TBPI patients, an effect we termed long afferent facilitation or LAF. LAF positively correlated with results in the Central Sensitization Inventory and in the Disabilities Arm, Shoulder, and Hand questionnaire.

**Conclusion:** These results point to the existence of an inhibitory regulation system between the representations of the face and the hand that seems to be suppressed in TBPI and correlates with pain. Moreover, brain changes arising from TBPI are not restricted to the hemisphere contralateral to the injured limb, but extend to both hemispheres.

## 1 Introduction

The classic description of sensorimotor cortex somatotopic organization suggests that motor actions involving different body regions are produced through the combined activation of clearly designated, independent cortical areas (Gross, 2007). However, this classic model of somatotopic organization, in which there is a precisely ordered representation of specific muscles or bodily movements in the primary cortex, has largely been discredited. (Donoghue, Leibovic, and Sanes, 1992; Park et al., 2001; Sanes and Schieber, 2001; Schieber, 2001; Graziano and Aflalo, 2007). For example, it has been shown that the electrical activation of specific cortical regions in primates generates movements, such as reaching, grasping, defending, and hand to mouth movements, that involve different body parts (Graziano, 2006). In respect of the face and hand, they have neighboring representations in the cortex although they are anatomically distant, and work very closely together in many everyday activities such as self-feeding and communicative manual gestures during speech (Gentilucci and Dalla Volta, 2008; Vainio, 2019). Moreover, interactions between the motor, the somatosensory, and the sensorimotor functions of the hand and the face have been observed in a number of different experimental contexts (Salmelin and Sams, 2002; Tanosaki et al., 2003; Higginbotham, Isaak, and Domingue, 2008; Desmurget et al., 2014). For instance, a repetitive sensory stimulus to the tip of the index finger resulted in an improvement in a two-point discrimination task not only at the stimulated finger, but also in the unstimulated ipsilateral face of the subject (Muret et al., 2014; Muret et al., 2016), indicating that passive stimulation of one body part can affect touch perception at a distant part of the body. The abundance of sensorimotor interactions between the hand and face and the magnitude of their representations could indicate anatomical correlates across their representation in the cortex, although evidence in this direction is still limited, and histological studies with monkeys have described the existence of a well-defined myelin septum between the areas of hand and face representation in area 3b of the somatosensory cortex (Jain, Catania, and Kaas, 1998), suggesting an anatomical separation between these two regions. However, other studies using marker injections and tracking techniques have identified fibers that project between the sensory and motor representations of the hand and face (Huntley and Jones, 1991; Fang, Jain, and Kaas, 2002).

The structural and functional proximity of hand and face representations is also evidenced after peripheral lesions (Elbert et al., 1997). Classically, it is described that cortical reorganization occurs with an expansion or displacement of the cortical representation of one area into neighboring regions originally occupied by afferent neurons from the injured body segment. The proximity of the representations of the face and the hand in the primary sensorimotor cortex favors these type of plasticity processes. Thus, the amputation of the hand, for example, induces the displacement of the cortical representation of the face towards the original hand representation area (Ramachandran, 1993; Flor et al., 1995; Pascual-Leone et al., 1996; Weiss et al., 2004; Raffin et al., 2016). Plasticity in this case may be accompanied by the sensation of the phantom hand upon stimulation of the face (Halligan et al., 1993; Ramachandran, 1993). Referred sensation is not, however, restricted to stimulation of neighboring regions, since it has also been described in upper limb amputees after touch applied to distant regions such as the feet, chest, or the opposite hand (Knetch et al., 1996; Grüsser et al., 2004). Thus, the stability of cortical representations, the cortical plasticity mechanisms and their limits, and the multiple factors involved in such processes have been the topic of intense discussion recently (Makin and Bensmaia, 2017; Makin and Flor, 2020).

In this context, traumatic brachial plexus injury (TBPI) presents itself as a suitable model for the study of the reorganization of cortical representations. The vulnerability of the brachial plexus to injury results from a number of factors, including its extension, its superficial position and its relatively lack of muscular or bone protection (Ferrante, 2004; Flores, 2006). In most cases, TBPI affects young male adults and is caused by closed trauma, mainly in automobile or motorcycle accidents leading to nerve rupture, avulsion at the spinal cord level, or significant nerve stretching without rupture (Moran, Steinmann, and Shin, 2005). Despite its very consistent epidemiological characteristics, TBPI engenders a great variability of clinical presentations. Factors such as injury mechanism, extension, the presence of pain, concomitant injuries, and the quality of medical and hospital care can influence treatment outcomes (Giuffre et al., 2010; Flores, 2011; Franzblau et al., 2015). Beyond sensorimotor loss, the appropriate management of pain is one of the biggest challenges for the health team involved in TBPI care (Santana et al., 2016). The estimates of the incidence of chronic pain after TPBI vary in the literature, with values often above 50%, and reaching 95% in some studies (Waikakul, Waikakul, and Pausawasdi, 2000; Flores, 2006; Vannier et al., 2008; Ciaramitaro et al., 2010; Giuffre et al., 2010; Jain et al., 2012). Furthermore, in TBPI patients, the upper limb not affected by the injury also undergoes significant changes. TBPI patients submitted to a thorough evaluation of tactile sensitivity with monofilaments on both limbs showed an increased sensitivity threshold in the uninjured side when compared to control participants (Ramalho et al., 2019). Motor synergies involving the upper limb and trunk, investigated through kinematic recording, were also found to be altered on the uninjured side in patients with TBPI (Souza et al., 2021). It is also clear that TBPI and its surgical reconstructions are capable of triggering cortical reorganization (Narakas, 1984; Mano et al., 1995; Malessy et al., 2003; Yoshikawa et al., 2012; Qiu et al., 2014). These plastic changes seem to be persistent (Hua et al., 2013), and correlate with the patients’ functional improvement (Kakinoki et al., 2017). Fraiman et al. (2016) observed a bilateral reduction in the M1 intrinsic activity responsible for upper limb synergies in TBPI patients, whereas facial functional connections in the M1 were preserved, suggesting that cortical changes presented by TBPI patients are bilateral but specific to the body part most directly affected by the injury. Moreover, these plastic changes seem to extend beyond the sensorimotor network and encompass higher-order cognitive networks such as the salience network and the default mode network (Bhat et al., 2017), also affecting sensorimotor interhemispheric connectivity and intra- and interhemispheric thalamic nuclei connectivity. For instance, in an fMRI study, patients with TBPI displayed weakened functional connectivity between hand grasp related areas and the supplementary motor area and multiple brain regions associated with motor processing or information integration (Yechen Lu et al., 2016).

A deeper investigation of the interaction between somatosensory and motor systems is, therefore, key to understanding cortical plasticity dynamics. To that end, the afferent inhibition technique stands out as a simple way to assess sensorimotor integration, and thus, a useful technique for assessing situations in which the uncoupling of sensory and motor information is observed (Bikmullina et al., 2009; Ferreri et al., 2012; Rossini et al., 2015). Tokimura et al. (2000) first described a TMS induced MEP reduction or inhibition in a muscle of interest when the magnetic pulse is preceded by an electrical stimulus applied to the skin. When the inter-stimulus interval (ISI) is less than 50 ms, the so-called short afferent inhibition (SAI) occurs; When longer intervals are used the phenomenon is known as long afferent inhibition (LAI) (Chen, Corwell, and Hallett, 1999). SAI has been very well described in cases in which the electrical stimulation is applied close to the target muscle. For example, there is a strong inhibition in hand muscles responses after the stimulation of a hand nerve or of the tip of the index finger (Chen, Corwell, and Hallett, 1999; Tokimura et al., 2000; Di Lazzaro et al., 2002; Sailer et al., 2002; Di Lazzaro et al., 2005; Helmich et al., 2005; Kukaswadia et al., 2005; Tamburin et al., 2005; Bikmullina et al., 2009; Asmussen et al., 2013; Lapole and Tindel, 2014). SAI has also been demonstrated between different regions of the upper limb, such as between the skin of the hand and the forearm and arm muscles (Helmich et al., 2005; Tamburin et al., 2005; Bikmullina et al., 2009), and even between an electrical stimulus in one hand and a muscle in the contralateral hand (Ruddy et al., 2016). Ramalho et al. (2022) investigated sensorimotor interactions between the face and hand, hypothesizing that if face and hand representations are functionally coupled, then the electrical stimulation of the face would inhibit hand muscle motor responses. They found that the delivery of a peripheral electrical stimulus to either the skin over the right upper lip or the right cheek inhibited muscular activity from the right first dorsal interosseous (Ramalho et al., 2022). These findings provided the first evidence of face-to-hand afferent inhibition.

In this study, TBPI was chosen as a model to investigate plasticity in face and hand cortical sensorimotor circuits. The aim was to investigate changes in hand and in face-to-hand sensorimotor integration in TBPI patients using the SAI and LAI afferent inhibition paradigm, assessing the hemispheres contralateral both to the injured and the uninjured limb. If AI could be induced in the hand of TBPI patients assessed on the injured side, preserved hand sensorimotor integration would be demonstrated. We hypothesized that if face-to-hand AI in TBPI patients assessed on the injured side was altered, it could indicate that changes in the cortical representations of the face and the hand caused by the TBPI can affect the sensorimotor integration between the face and the hand. Since TBPI also causes altered function in the uninjured limb and bilateral brain changes, face-to-hand AI should also be altered in TBPI patients assessed on the uninjured side.

## 2 Materials and Methods

### 2.1 Participants

Patients with a diagnosis of unilateral TBPI were recruited using a digital database developed at the Laboratory of Neurosciences and Rehabilitation (LabNeR) (Patroclo et al., 2019) and stored on the Neuroscience Experiment System (NES) platform (Ruiz-Olazar et al., 2022). This database consists of a collection of clinical and neurophysiological information from adult patients with TBPI treated at the Institute of Neurology Deolindo Couto (INDC) at the Federal University of Rio de Janeiro (UFRJ), or at the National Institute of Traumatology and Orthopedics Jamil Haddad (INTO). Recruitment took place between September 2019 and March 2020 and a selection based on clinical data produced a contact list with 23 patients. Patients were excluded if they had a history of severe traumatic brain injury, prolonged loss of consciousness during the accident that led to the injury, long-term use of drugs affecting the CNS or the presence of metal implants near stimulation sites. After systematic attempts to contact patients on the preliminary list, 10 agreed to participate in the experiments after the confirmation of the inclusion and exclusion criteria explained below. Nine patients actually participated in the experiments, forming the TBPI group. Although the number of participants was small, it was similar to that in previous studies with TMS (Batista e Sá et al., 2015; Mercier et al., 2006; Tokimura et al., 2000), and it was adapted by the need to comply with social distancing rules in force during the COVID-19 pandemic. In addition, 18 healthy volunteers from the UFRJ community (students and employees) were recruited to form a control group (CG). For the selection process, the inclusion criteria for all participants were: age between 18 years and 55 years; any gender; preserved communication ability; tolerance to remain seated for at least 2 hours; and, for the TBPI group: a clinical diagnosis of complete or incomplete unilateral brachial plexus traumatic injury. The exclusion criteria were a history of psychiatric illness, including substance abuse, or cognitive impairment; a history of diseases and/or sequelae of the central nervous system (CNS) or peripheral nervous system (PNS); a history of chronic pain previous to the TBPI (for the TBPI group) or any report of chronic pain (for the CG); and answering YES to any question in the safety screening questionnaire for TMS application (adapted from Rossi et al., 2011). All participants underwent the experimental protocol at LABNER between May 2019 and January 2022, and those in the TBPI group underwent clinical and pain assessment at LABNER or INTO between January 2019 and May 2020. The TMS protocol and the TBPI database enrollment were approved by the research ethics committee of INDC-UFRJ (registered numbers: 2.411.426 and 2.087.610, respectively) and all participants gave written informed consent to participate in the experiments. Previous research with TBPI patients has demonstrated changes in sensation (Ramalho et al., 2019) and kinematics (Souza et al., 2021) in the uninjured limb, and has also shown that the sensorimotor cortex related to the uninjured side undergoes plasticity (Hsieh et al., 2002; Liu et al., 2013; Fraiman et al., 2016; Rangel et al., 2021), which means not only that TBPI patients cannot be used as controls for themselves, but also that the AI protocol can be applied to the uninjured limb in TPBI participants to examine changes in plasticity. This is of particular importance in relation to the face-hand sensorimotor motor circuits, as these cannot be assessed in the injured limb of patients with complete injury where there is a complete absence of sensory and/or motor function in the hand. Therefore, the TBPI participants were subdivided into two groups based on their clinical characteristics and diagnosis, with those with at least partial sensorimotor function in the hand (i.e., incomplete injury with an upper trunk or extended upper trunk injury diagnosis) allocated to the *TBPI assessed on the injured side* (TBPI-I) group, while those without any hand sensorimotor function (i.e., mostly complete injury diagnosis) were allocated to the *TBPI assessed on the uninjured side* (TBPI-UI) group. Thus, the final research paradigm was composed of three experimental groups: TPBI group, divided into TBPI-I and TBPI-UI, plus the Control Group (Figure 1).

**Figure 1.**
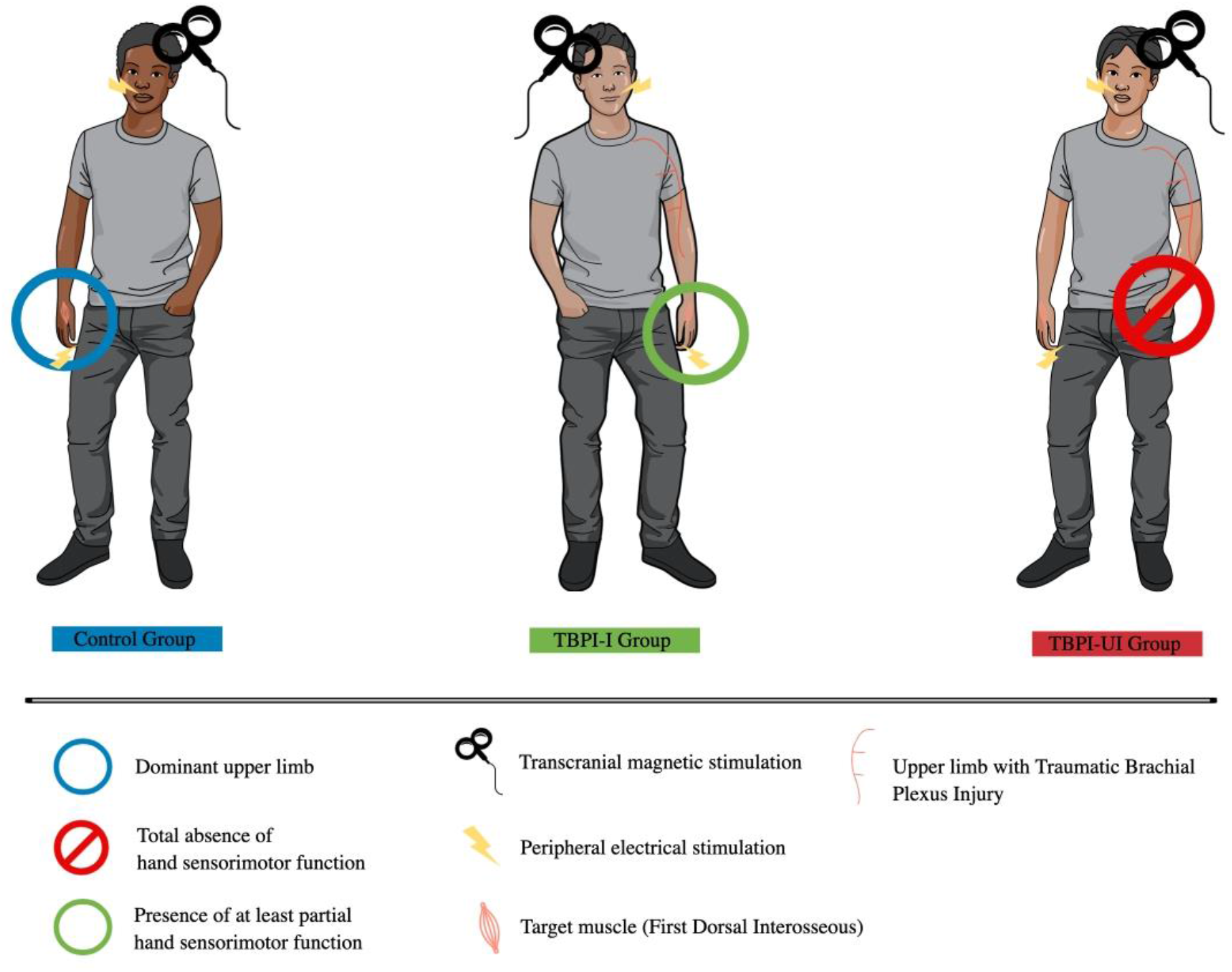
Experimental groups. Control group (in blue, on the left) was assessed on the dominant upper limb. The TBPI-I group (in green, center) was formed by participants with at least partial sensorimotor function in the hand of the injured side, which allowed it to be evaluated using the afferent inhibition protocol. The TBPI-UI group (in red, on the right) was formed by participants with total absence of sensorimotor function in the hand, thus they were assessed on the uninjured side. Peripheral electrical stimulation was applied either on the tip of the ipsilateral index finger or above the upper limb. Transcranial magnetic stimulation was applied over the scalp contralateral to the assessed limb.

### 2.2 Clinical assessment and pain evaluation

All TBPI participants were clinically evaluated through instruments developed by the LABNER research group (unified entry evaluation, unified follow-up evaluation, and surgical evaluation), which are described in detail elsewhere (Patroclo et al., 2019). The information that was collected and assessed included demographic data, injury details, clinical history, previous pathological history, physical examination data (inspection findings, muscle trophism, range of motion, strength, and superficial and deep sensation), previous treatments and surgical procedures. The evaluation also included the Disabilities Arm, Shoulder, and Hand (DASH) questionnaire (Orfale et al., 2005) to assess upper limb functionality, and the post-traumatic stress disorder checklist: civilian version (PCL-C) (Bringhenti, Luft, and de Oliveira, 2010), to screen for post-traumatic stress disorder.

Additionally, a detailed pain evaluation was performed for each TBPI participant through specific instruments, namely: a visual analog scale (VAS) to determine the intensity of pain (Downie et al., 1978); the Brief Pain Inventory (BPI), to quantify how pain interferes in various aspects of daily life (Toledo, 2008; Ferreira et al., 2011); a body pain map, part of the BPI, to register pain location (adapted from Margolis, Tait, and Krause, 1986; Toledo, 2008; Ferreira et al., 2011); the Douleur Neuropathique 4 Questions (DN-4), a questionnaire capable of identifying and classifying pain of neuropathic origin (Santos et al., 2010); the McGill questionnaire, which allows for a qualitative assessment of pain (Pimenta and Teixeira, 1996); and finally, the Central Sensitization Inventory (CSI), to assess the degree of central sensitization (Caumo et al., 2017). The approximate time for complete clinical assessment was approximately 2 hours. Furthermore, on the day of the experimental protocol, upon arrival at LABNER, all participants completed the Edinburgh Handedness Inventory (Oldfield, 1971), the Mini Mental State Examination (MMSE) (Brucki et al., 2003) and the safety screening questionnaire for the application of TMS (adapted from Rossi et al., 2011).

### 2.3 Transcranial magnetic stimulation and electromyography

TMS was applied to the scalp contralateral to the side of the studied limb: the dominant side for the CG, the uninjured side for the TPBI-UI group, and the injured side for the TPBI-I group. TMS was performed with a Magstim 200^2^ stimulator (The Magstim Company, Carmarthenshire, UK) using a figure-of-eight coil with 70 mm internal diameter for each wing, positioned tangentially to the skull over M1, with the coil handle pointing backwards and an angulation of 45° from the midline. A neuronavigation system (InVesalius Navigator 3.1.1 - 3 Space TM Fastrack® - Polhemus Isotrack II) was used to ensure the accuracy of the TMS coil positioning on the participants’ scalps and the consistency of the stimulus applied during the experimental sessions (Souza et al., 2018).

Motor output was recorded with surface EMG on the contralateral first dorsal interosseous muscle (FDI). Adhesive surface electrodes were used for the EMG in a bipolar configuration (Ag/AgCl in solid gel, 28 × 20 mm, with 20 mm distance between electrodes) (Neuroline 715, Ambu, Copenhagen, Denmark). Electrodes were positioned on the FDI muscle belly and on the styloid process of the ulna ipsilateral to the recording site (ground electrode). The EMG signal was collected at a 2000 samples/second sampling rate, with a gain of 1000 x, filtered through a 2-pole Butterworth bandpass filter with a frequency range of 20-500 Hz”, and digitized by a recording platform consisting of a CED 1902 amplifier and a CED Power 1401 data acquisition unit (both Cambridge Electronic Design Limited, Cambridge UK). The EMG signal was then processed with Signal software version 6.05 (Cambridge Electronic Design Limited, Cambridge UK) and was stored on a computer for further data analysis.

During the experimental sessions, participants were seated in a comfortable chair with arm support, maintaining approximately 90° of elbow flexion with the wrist in a neutral position. Participants were instructed to keep the upper limb relaxed, to avoid talking during the experiments, to keep both feet flat on the floor, and to remain awake with eyes open. The EMG signal was constantly visually monitored to ensure that the target muscle was completely relaxed. If muscle activity was detected, the participant was verbally instructed to relax.

The following measurements were obtained:

a. FDI hot spot: the optimal point for stimulation of the FDI muscle was defined as the region that, when stimulated, generated stable MEPs with larger peak-to-peak amplitudes, reproducible for at least five consecutively applied stimuli.
b. Resting motor threshold (rMT): defined as the minimal TMS stimulation intensity required to obtain MEPs with at least 50 μV peak-to-peak amplitude in at least 5 out of 10 consecutively applied stimuli.
c. Motor threshold to elicit 1 mV responses (1 mV/MT): defined as the minimum stimulation intensity required to obtain MEPs with at least 1 mV peak-to-peak amplitude in at least 5 out of 10 consecutively applied stimuli. The 1 mV target was chosen to obtain MEPs sufficiently large to allow the observation of the inhibition phenomenon (Turco et al., 2018; Ramalho et al., 2022).

### 2.4 Peripheral electrical stimulation

Peripheral electrical stimulation was applied by a constant current stimulator (STMISOLA, BIOPAC Systems Inc., USA) and consisted of square wave pulses with short duration (200 μs) and intensity determined by the participant’s perception of a non-painful sensory perception. Stimulation electrodes were positioned in a bipolar configuration on the palmar side of the index fingertip or on the region above the upper lip next to the lip philtrum, placed side by side with a 1 cm separation between each center. Adhesive surface electrodes (Ag/AgCl in solid gel 28 × 20 mm, Neuroline 715, Ambu, Copenhagen, Denmark) were adapted to obtain an approximate size of 15 × 15 mm.

The following measurements were obtained:

a. Peripheral electrical threshold (pET) for the finger and the face: determined according to the participant’s perception so that the stimulus would produce a non-painful sensory perception, and defined as the lowest stimulation intensity perceptible in 10 consecutive stimuli.
b. Peripheral electrical stimulation intensity (pESI) for the finger and the face: the electrical stimulation intensity applied during the experiments was set at 3xpET for the finger and 2xpET for the face, thus ensuring a stimulus below the nociceptive threshold. This choice was based on AI protocols typically used (Tokimura et al., 2000; Tamburin et al., 2002; Tamburin et al., 2005; Bikmullina et al., 2009; Tamè et al., 2015) and on previous experiments from the LABNER research group (Ramalho et al., 2022), showing that a stimulation intensity of 3xpET for the face was painful for most participants.

### 2.5 Experimental protocol

The peripheral electrical stimulation was applied on the skin above the upper lip or on the index fingertip succeeded by a single TMS pulse over the FDI hot spot in M1, with different inter-stimulus intervals (ISI) to evoke short and long AI. Single TMS pulses without electrical stimulation were used as the control condition. ISIs were 15, 25, 35, 45, 55 and 65 ms for SAI and 100, 200, 300 and 400 ms for LAI. Significant FDI SAI has been reported with ISIs between 25 and 50 ms and face-hand inhibition with ISIs between 45 and 65 ms (Helmich et al., 2005; Tamburin et al., 2005; Bikmullina et al., 2009; Ramalho et al., 2022). ISIs were then chosen based on the premise that SAI and LAI would occur at longer intervals for TPBI participants, considering the nerve injury. The order of execution of the four blocks was determined by a drawing for each participant: SAI and LAI between the index fingertip and the FDI hand muscle, and SAI and LAI between the face and the FDI hand muscle. For each block, 14 paired electrocutaneous and TMS stimuli were applied for each ISI with an additional 14 single TMS pulses for the control condition. Different ISI stimuli were presented randomly, with intervals of 2.5 to 4 s between each paired or single TMS pulse. SAI blocks had a total of 98 pulses and LAI blocks a total of 70 pulses. For each block there was a brief pause of about 1 minute after every 49 pulses (SAI) or 35 pulses (LAI). Average duration of the entire experiment session was around 2 hours and 30 minutes.

### 2.6 Data processing and statistical analysis

For signal analysis, the peak-to-peak amplitude of each MEP was measured in real time (Signal software, Cambridge Electronic Design Limited, Cambridge, UK). To ensure data quality, trials with clear evidence of muscle contraction or with EMG artifacts that prevented MEP measurement were excluded. Trials in which the peak-to-peak amplitudes were too small or in which there was an absence of MEP were accounted for, but not excluded. This decision was made because it would not be possible to determine whether the absence of MEPs occurred due to a modulatory effect of the electrostimulation, a TMS accommodation effect causing progressively smaller MEPs, or even due to some undetermined technical issue. To detect possible atypical results, an outlier exclusion criterion was applied. Trials with a peak-to-peak amplitude of < Q1-1.5xIQR or > Q3-1.5xIQR for each condition of each block for each participant were excluded. Thus, the total number of lost trials was counted. The maximum losses permitted per block per participant was 30% of the total trials. When losses exceeded the stipulated maximum, the participant was excluded from the block in question. Furthermore, participants with mean peak-to-peak amplitude MEPs lower than 0.3 mV for the control condition (single TMS pulse) were also excluded, since it would be difficult to observe the inhibition phenomenon with MEPs this size. Finally, a visual inspection of the temporal MEP sequence for each participant was performed, which made it evident that for the first two CG participants there was a malfunction of the electrostimulation equipment, which failed to elicit the desired stimuli during entire experimental blocks. These data could not be used in subsequent analyses, so the two participants were excluded. Table S.1 and figure S.1 in the Supplementary Material show the exclusion process details and the data distribution before and after outlier removals and exclusion process. Figures 2 and 3 show the final number of participants per experimental block.

**Figure 2.**
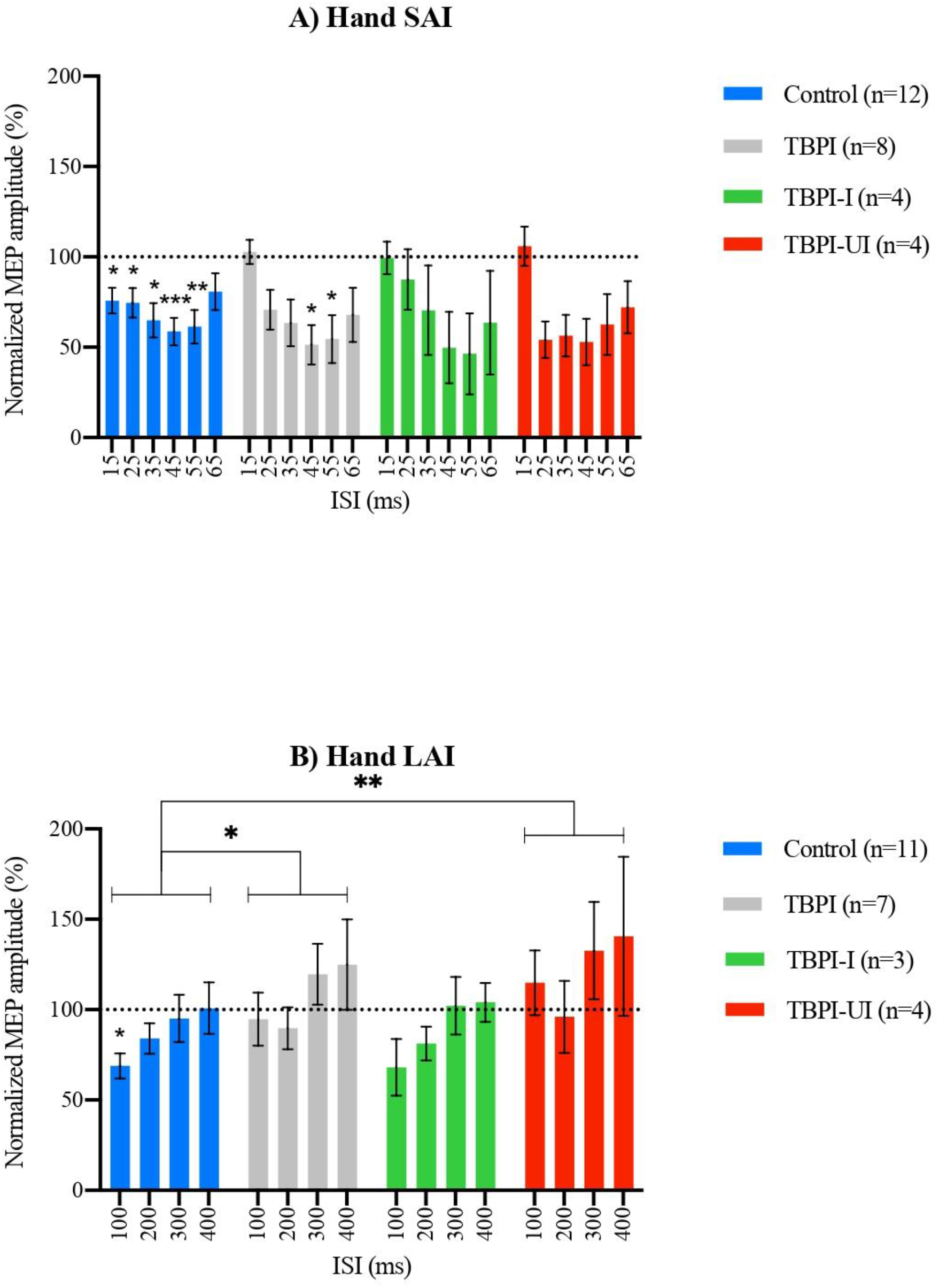
Hand SAI and LAI results. Peripheral electrical stimulation applied on the tip of the index finger followed by contralateral transcranial magnetic stimulation over the first dorsal interosseous hot spot. Different inter-stimulus intervals were applied so as to include short afferent inhibition (SAI) intervals (A) and long afferent inhibition (LAI) intervals (B). Group mean motor evoked potential amplitudes for each inter-stimulus interval normalized by mean amplitudes in the control condition (transcranial magnetic stimulation without previous peripheral electrical stimulation). Control Group (blue); TBPI group, all TBPI participants (gray); TBPI-I group, TBPI patients assessed on the injured side (green), and TBPI-UI, TBPI patients assessed on the uninjured side (red). Bars represent the standard error of the mean. Values below the dotted line at 100% indicate an inhibition effect, values above the dotted line indicate a facilitation effect. Repeated measures one-way ANOVA and Dunnett’s post-test were performed for intragroup analysis (* above bars). Two-way ANOVAs and Šidák’s or Tukey’s post-test were used for intergroup comparisons (* above brackets). * = p ≤ 0,05; ** = p ≤ 0,01; *** = p ≤ 0,001.

**Figure 3.**
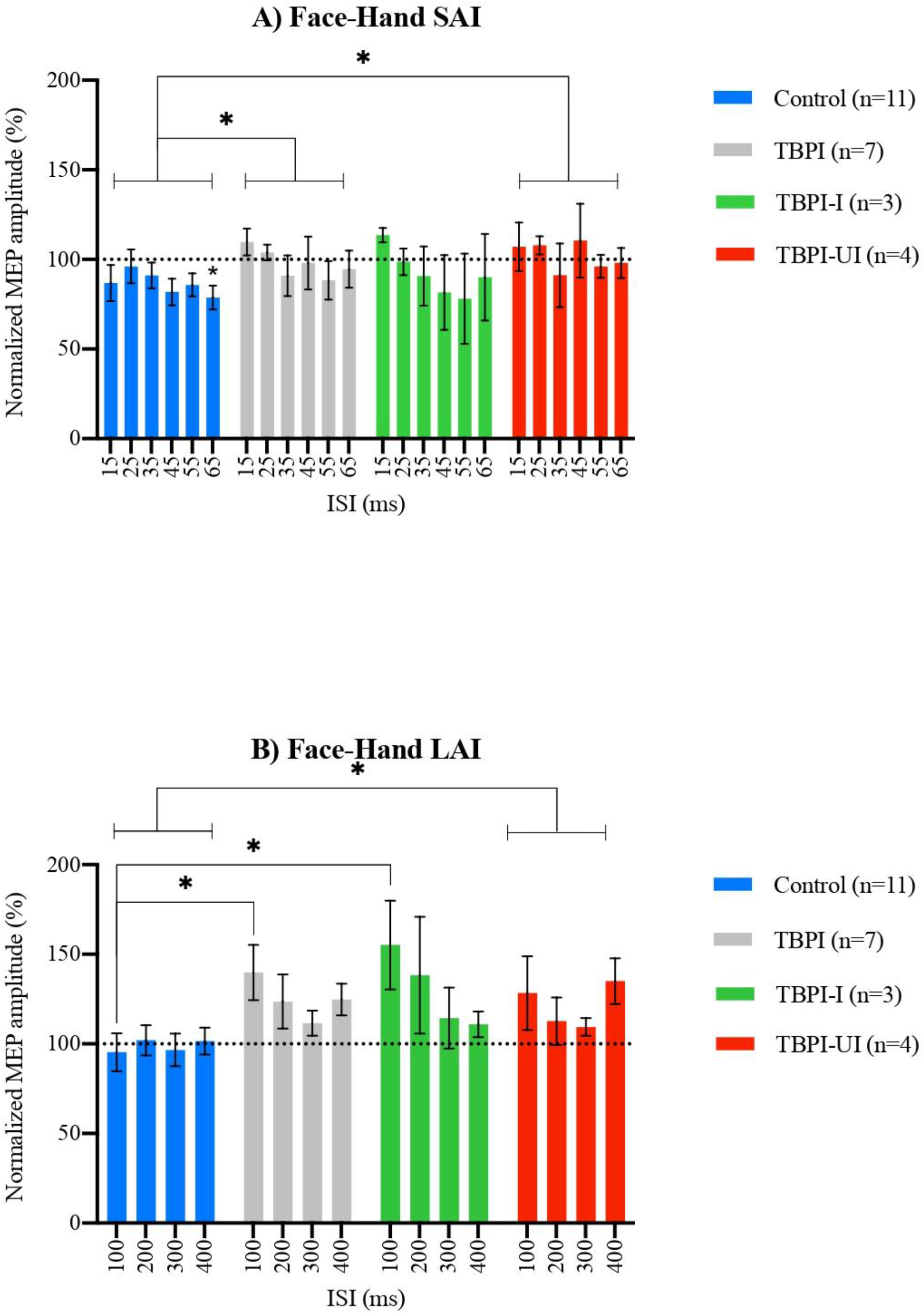
Face-to-hand SAI and LAI results. Peripheral electrical stimulation applied on the face, above the upper lip, followed by contralateral transcranial magnetic stimulation over the first dorsal interosseous hot spot. Different inter-stimulus intervals were applied so as to include short afferent inhibition (SAI) intervals (A) and long afferent inhibition (LAI) intervals (B). Group mean motor evoked potential amplitudes for each inter-stimulus interval normalized by mean amplitudes in the control condition (transcranial magnetic stimulation without previous peripheral electrical stimulation). Control Group (blue); TBPI group, all traumatic brachial plexus injury participants (gray); TBPI-I group, TBPI patients assessed on the injured side (green); and TBPI-UI, TBPI patients assessed on the uninjured side (red). Bars represent the standard error of the mean. Values below the dotted line at 100% indicate an inhibition effect, values above the dotted line indicate a facilitation effect. Repeated measures one-way ANOVA and Dunnett’s post-test were performed for intragroup analysis (* above bars). Two-way ANOVAs and Šidák’s or Tukey’s post-test were used for intergroup comparisons (* above brackets). * = p ≤ 0,05; ** = p ≤ 0,01; *** = p ≤ 0,001.

For statistical analysis, mean MEP peak-to-peak amplitudes (mV) were obtained for every condition in each block for each participant. Mean MEP amplitudes for different ISIs were normalized by the mean MEP amplitude for the control condition for each participant individually. Next, mean MEP peak-to-peak amplitudes (mV) and mean normalized MEP amplitudes (%) for each ISI were obtained for each block for the different groups. Normalized data was used for the statistical analysis. Therefore, when interpreting results, values below 100% indicate inhibition and values above 100% indicate facilitation. The Shapiro-Wilk test was used to verify whether data could be described by a normal distribution. For within group comparisons repeated measures one-way ANOVA and Dunnett’s multiple comparisons post-test were performed to compare normalized MEP amplitude means in each ISI with the control condition (single TMS pulse = 100%). For intergroup comparisons, two-way ANOVA were performed using ISI and groups as factors. This was first done comparing groups in pairs (CG x TBPI; CG x TBPI-I; CG x TBPI-UI; TBPI-I x TBPI-UI) using Šidák’s multiple comparisons test as the post hoc analysis; and then comparing the three groups (CG x TBPI-I x TBPI-UI) using Tukey’s multiple comparisons test as post hoc. To compare experimental parameters (mean MEP amplitude for the control condition, 1mV/MT, finger and face pET and pESI) between different groups, ordinary one-way ANOVA using group as factor and Tukey’s multiple comparisons test were used. Finally, for the TBPI group, the interval with the greatest modulatory effect (inhibition or facilitation) was selected for each block and individual normalized values for the selected ISI were used to assess if there was a linear relationship with demographic and clinical data (age, time between injury and surgery, time between surgery and participation on the experiment), questionnaire results (Edinburgh Handedness Inventory, MMSE, DASH and PCL-C scores) and pain scores (VAS, DN4, BPI, CSI, McGill scores). For this, Pearson correlation coefficients were computed. Statistical significance was defined at 5%. All data were analyzed with Prism 9 software (GraphPad Software, Inc., California, USA). For graphic representation, normalized MEP amplitudes for every ISI are presented with mean and standard error for each group.

## 3 Results

### 3.1 General Characteristics, clinical assessment and pain evaluation

General social characteristics and the results for the Edinburgh Laterality Inventory and the MMSE for all participants are shown in Table 1. Five TBPI patients were evaluated on the injured side (TBPI-I Group) and four were evaluated on the uninjured side (TBPI-UI). Information about TBPI history, diagnosis, upper limb functionality, physical exam and previous treatments are displayed in Table 2 and information regarding the pain characteristics of the TBPI patients is presented in Table 3.

**Table 1.** Age and scores presented as mean (maximum – minimum). TBPI-I, traumatic brachial plexus injury participants assessed on the injured side; TBPI-UI, traumatic brachial plexus injury participants assessed on the uninjured side; M, male; F, female; RH, right-handed; L, left-handed; W, white; B, black; ASB, Asian-Brazilian; EHI, Edinburgh Handedness Inventory score, in which scores of < -40 indicate left dominance and scores > 40 indicate right dominance and scores between -40 and 40 indicate ambidexterity; MMSE, Mini-Mental State Examination, scores ranging from 0 to 30, with scores above 27 indicating normality of cognitive function for people with at least four years of schooling and scores above 17 indicating normality for people with less than four years of schooling.

| Demographic and social characteristics of participants |  |  |  |  |
| --- | --- | --- | --- | --- |
|  | Control Group (n=18) | TBPI-I Group (n=5) | TBPI-UI Group (n=4) | All (n=27) |
| Age | 28 (20 – 54) | 36 (25 – 47) | 28.5 (24 – 54) | 28 (20 – 54) |
| Gender | 15 M / 3 F | 5 M | 4 M | 24 M / 3 F |
| Laterality | 16 RH / 2 LH | 5 RH / 0LH | 3 RH / 1 LH | 24 RH / 3 LH |
| EHI | 70.7 (-79.0 – 90.0) | 79.0 (74.2 – 94.9) | 54.9 (-68.4 – 63.2) | 71.4 (-79.0 – 94.9) |
| MMSE | 29 (26 – 30) | 28 (23 – 29) | 28 (28 – 30) | 29 (23 – 30) |
| Ethnicity/race | 13 W / 4B / 1 ASB | 3 W / 2 B | 3 B / 1 W | 17 W / 9 B / 1 ASB |

**Table 2.** R, right; L, left; y, years; m, months; N, no; Y, yes; BP exploration, brachial plexus, exploration; Oberlin, transfer of an ulnar fascicle to the biceps branch of the musculocutaneous nerve; A/SS, transfer of the accessory nerve to the suprascapular nerve; T/A, axillary nerve neurotization by a triceps motor branch; Neurotization S/P/A, neurotization with sural, phrenic and accessory nerve grafts; P/MC, Phrenic nerve transfer to the musculocutaneous nerve; A/MC, accessory nerve transfer to the musculocutaneous nerve; ↑, increased, ↓, decreased, Ø, absent C3, C4, C5, C6, C7, C8, T1, T2: matching dermatomes; DASH, Disabilities Arm, Shoulder and Hand questionnaire score, ranging from 0 (maximum functionality) to 100 (maximum disability); PCL-C, Post-Traumatic Stress Disorder Checklist Civilian Version total scores, ranging from 17 to 85, where a 44 points score is indicative of PTSD. However, the diagnosis of post-traumatic stress must be made according to the diagnostic rules of DSM-5 (Diagnostic and Statistical Manual of Mental Disorders), by a specialized professional.

| Clinical characteristics of the TBPI participants |  |  |  |  |  |  |  |  |  |
| --- | --- | --- | --- | --- | --- | --- | --- | --- | --- |
|  | TBPI-I1 | TBPI-I2 | TBPI-I3 | TBPI-I4 | TBPI-I5 | TBPI-UI-1 | TBPI-UI2 | TBPI-UI3 | TBPI-UI4 |
| Handedness | R | R | R | R | R | R | R | L | R |
| Injured side | L | L | R | R | R | L | L | L | L |
| Age range | 35-40 | 30-35 | 45-50 | 45-50 | 25-30 | 20-25 | 25-30 | 25-30 | 50-54 |
| Diagnosis | Extended upper trunk | Extended upper trunk | Upper trunk | Extended upper trunk | Extended upper trunk | Complete | Complete | Upper trunk | Complete |
| Cause | Motorcycle accident | Motorcycle accident | Motorcycle accident | Motorcycle accident | Motorcycle accident | Motorcycle accident | Motorcycle accident | Motorcycle accident | Motorcycle accident |
| Time since injury | 4y 10m | 5y 10m | 1y 7m | 5y 2m | 4y 6m | 1y 5m | 12y 1m | 4y 1m | 3y 5m |
| Surgery | BP Exploration | Oberlin ± A/SS | Oberlin ± A/SS ± T/A | Oberlin | Oberlin | Neurotization S/P/A | - | P/MC | A/MC |
| Time injury-surgery | 10m | 5m | 3m | 11m | 1y 1m | 7m | - | 4m | 6m |
| Orthopedic surgery | clavicle, arm | clavicle | leg | arm, forearm, thigh, knee | - | clavicle | - | arm, hand | cervical vertebra |
| Physical therapy | N | N | Y | Y | N | Y | N | Y | Y |
| Tactile sensitivity | ↑C5, T3 ± ↓C6-7-8 | ↓C3, C5, C7-8 ± ØC4, C6 | ØC4-T2 | ↑C4-8 ± ↓T1-3 | ↑C4 ± ↓C5 | ↓C5-6, T1 ± ØC6-8 | ØC5-T1 | ↑C5 ± ↓C6-T1 | ↓C3-5 ± ØC6-8 |
| Pain sensitivity | ↑C3-4, T1, T3 ± ↓T2 ± ØC5-8 | ↓C8 ± ØC4-7 | ØC6-8 | ↑C4 ± ↓C5, C8-T1 ± ØC6-7, T2-3 | ↓C4 ± ØC5 | ↓C4 ± ØC5-T1 | ØC5-T1 | ↓C6-8 ± ØC5, T1 | ↓C4-5, T1-2 ± ØC6-8 |
| Strength | ↓shoulder, elbow | ↓shoulder | ↓shoulder | ↓shoulder, elbow, hand | ↓shoulder, elbow | Øshoulder, elbow, hand | Øshoulder, elbow, hand | ↓shoulder, elbow, Øhand | Øshoulder, elbow, hand |
| DASH | 33.33 | 25 | 40.52 | 31.67 | 40.83 | 19.64 | 22.5 | 49.17 | 68.33 |
| PCL-C | 27 | 29 | 18 | 29 | 32 | 37 | 19 | 67 | 31 |

**Table 3.** TBPI-I, traumatic brachial plexus injury participants assessed on the injured side; TBPI-UI, traumatic brachial plexus injury participants assessed on the uninjured side; Intensity, assessed by the visual analog scale (VAS) for pain, ranging from 0 (no pain) to 10 (maximum pain intensity); m, months; Localization, pain sites defined by the body map of pain presented in item 2 of the brief pain inventory; DN4, Douleur Neuropathique 4 Questions score, ranging from 0 to 10, 4 being the cutoff point for defining neuropathic pain; CSI-A, part A Central Sensitization Inventory score, ranging from 0 to 100, with higher values indicating a higher degree of central sensitization; CSI-B, responses to part B of the Central Sensitization Inventory, which assesses whether the patient has ever been diagnosed with any of the diseases included in the central sensitization syndrome and, if so, the year of this diagnosis; McGill – SS, McGill Questionnaire sensory pain score; McGill – AS, McGill Questionnaire affective pain score on; McGill – ES, McGill Questionnaire evaluative pain score; McGill – MS, McGill Questionnaire miscellaneous pain score; McGill – PRI, McGill Questionnaire pain rating index obtained through the sum of the intensity values of the descriptors chosen, with a maximum value of 78; BPI, Brief Pain Inventory score, average of responses to item 9 of the questionnaire, which ranges from 0 to 10.

| Pain characteristics of the TBPI participants |  |  |  |  |  |  |  |  |  |
| --- | --- | --- | --- | --- | --- | --- | --- | --- | --- |
|  | TBPI-I1 | TBPI-I2 | TBPI-I3 | TBPI-I4 | TBPI-I5 | TBPI-UI1 | TBPI-UI2 | TBPI-UI3 | TBPI-UI4 |
| Intensity | 3 | 8 | 2 | 6 | 10 | - | 10 | 10 | 7 |
| Duration | >12m | >12m | >12m | >12m | >12m | - | >12m | >12m | >12m |
| Localization | 51 | 51 | 22 | 50, 34 | 4, 19, 35, 42, 51, 52 | - | 9, 35, 42, 51, 52 | 9, 22, 35, 42, 51, 52 | 4, 9, 19, 22, 35, 42, 51, 52 |
| DN4 | 3 | 6 | 5 | 3 | 3 | - | 5 | 8 | 7 |
| CSI-A | 30 | 9 | 33 | 17 | 20 | - | 6 | 41 | 37 |
| CSI-B | - | - | Cervical injury (2010) | - | - | - | - | - | Cervical injury (2016) |
| McGill – SS | 15 | 1 | 18 | 15 | 23 | - | 23 | 33 | 21 |
| McGill – AS | 6 | 0 | 6 | 5 | 7 | - | 13 | 12 | 7 |
| McGill – ES | 1 | 4 | 2 | 3 | 5 | - | 5 | 3 | 3 |
| McGill – MS | 4 | 0 | 6 | 7 | 5 | - | 11 | 14 | 5 |
| McGill – PRI | 26 | 5 | 32 | 30 | 40 | - | 52 | 62 | 36 |
| BPI | 3.71 | 1.57 | 1 | 2.57 | 5.71 | - | 6.86 | 8.29 | 4.86 |

### 3.2 Transcranial magnetic stimulation and electrocutaneous stimulation parameters

Transcranial magnetic stimulation and peripheral electrical stimulation parameters (mean <u>±</u> standard deviation) for each group are shown in Table 4. A one-way ANOVA was performed to compare the group effect for each parameter. No statistically significant difference was revealed for mean MEP amplitude in the control condition (F_2, 20_ = 0.1126, p= 0.894), for the 1mV/MT (F_2, 24_ = 0.1367, p= 0.873), for the face pET (F_2, 20_ = 1.729, p= 0.203) or the face pESI (F_2, 20_ = 1.611, p= 0.225). For the finger pET and pESI there was a significant difference between the Control and TBPI-I groups (F_2, 23_= 7.955, p= 0.002; F_2, 23_ = 7.538, p= 0.003) as evidenced by the post-hoc Tukey’s multiple comparisons test (p = 0.003, 95% C.I. = [-3.227; -0.6367]; p = 0.002, 95% C.I. = [-9.753; -2.050]).

**Table 4.** Results presented as mean ± standard deviation. Intergroup comparisons by ordinary one-way ANOVA and Tukey’s multiple comparisons test. TBPI-I, traumatic brachial plexus injury participants assessed on the injured side; TBPI-UI, traumatic brachial plexus injury participants assessed on the uninjured side; MEP amplitude (control), mean motor evoked potential amplitude in the control condition (transcranial magnetic stimulation without previous peripheral electrical stimulation); 1mV/MT, motor threshold to elicit 1 mV responses, presented as a percentage of the maximum power of the device; pET, peripheral electrical threshold for the finger and the face; pESI, peripheral electrical stimulation intensity for the finger and the face set at 3xpET for the finger and 2xpET for the face. * = p ≤ 0,05.

| Transcranial Magnetic Stimulation and Peripheral Electrical Stimulation Parameters |  |  |  |
| --- | --- | --- | --- |
|  | Control Group (n=18) | TBPI-I Group (n=5) | TBPI-UI Group (n=4) |
| Control MEP amplitude (mV) | 0.71 ± 0.42 | 0.73 ± 0.34 | 0.98 ± 0.47 |
| 1 mV/MT ( % ) | 52.17 ± 12.17 | 50.60 ± 9.02 | 54.50 ± 6.86 |
| Finger pET (mA) | 2.69 ± 0.82* | 4.62 ± 1.75* | 3.90 ± 0.55 |
| Face pET (mA) | 2.02 ± 0.99 | 2.96 ± 2.08 | 1.48 ± 0.51 |
| Finger pESI (mA) | 7.96 ± 2.37* | 13.86 ± 5.25* | 10.28 ± 1.84 |
| Face pESI (mA) | 4.46 ± 2.10 | 5.92 ± 4.16 | 2.85 ± 1.09 |

### 3.3 Short and long afferent inhibition in hand and face-hand sensorimotor circuits

Figure 2 shows the means and standard errors for normalized MEP amplitudes at SAI (Figure 2-A) and LAI (Figure 2-B) intervals after peripheral electrical stimulation of the index finger tip for each group. For the CG, inhibition was observed at all ISI with a maximum of 41.28% ± 7.57 at 45 ms. As for the TBPI group, inhibition was observed at 25, 35, 45, 55 and 65 ms, with maximum inhibition of 48.63% ± 10.91 at 45 ms. When the TBPI group was subdivided, TBPI-I displayed inhibition at 35, 45, 55 and 65 ms, with maximum inhibition of 53.62% ± 22.38 at 55 ms, and TBPI-UI showed inhibition at 25, 35, 45, 55 and 65 ms, with maximum inhibition of 47.05% ± 12.84 at 45 ms. Repeated measures one-way ANOVA for normalized MEP amplitude means at different ISIs within each group revealed a significant difference between the CG (F_3.131, 34.44_ = 4.171, p= 0.012) and the TBPI group (F_3.106, 21.74_ = 6.128, p= 0.003). The post-hoc Dunnett’s multiple comparisons test found significant inhibition at 15 ms (p = 0.025, 95% C.I. = [2.900; 45.47]), 25 ms (p = 0.043, 95% C.I. = [0.7259; 50.13]), 35 ms (p = 0.016, 95% C.I. = [6.376; 63.74]), 45 ms (p = 0.001, 95% C.I. = [18.43; 64.12]) and 55 ms (p = 0.007, 95% C.I. = [10.93; 66.39]) for the CG and at 45 ms (p = 0.012, 95% C.I. = [12.28; 84.97]) and 55 ms (p = 0.045, 95% C.I. = [1.099; 89.89]) for the TBPI group. There was no statistically significant difference in respect of SAI intervals for the hand when the TBPI were subdivided into TBPI-I (F_1.905, 5.715_ = 2.803, p= 0.143) and TBPI-UI (F_2.004, 6.011_ = 4.942, p= 0.054). To compare normalized MEP amplitude means at each ISI between the different groups, a two-way ANOVA using ISI and Groups as factors and the Tukey’s multiple comparisons test were used. For hand SAI, there was no main effect for Group when comparing groups in pairs (CG x TBPI: F_1, 108_ = 0.02387, p=0.878; CG x TBPI-I: F_1, 84_ = 0.0004983, p=0.982; CG x TBPI-UI: F_1, 84_ = 0.08343, p=0.773; TBPI-I x TBPI-UI: F_1, 36_ = 0.04558, p=0.832) or when comparing the three groups (CG x TBPI-I x TBPI-UI: F_2, 102_ = 0.03919, p=0.962).

For LAI intervals, the CG exhibited inhibition at 100 ms (31.10% ± 6.93) and 200 ms (15.99% ± 8.35). The TBPI group did not display inhibition but facilitation at 300 ms (19.60% ± 16.78). Once subdivided, the TBPI-I group showed inhibition at 100 ms (31.94% ± 15.67) and 200 ms (18.71% ± 9.35) but the TBPI-UI group showed facilitation at 300 ms (32.60% ± 26.95). RM one-way ANOVA showed no significant hand LAI for the CG (F_2.010, 20.10_ = 3.297, p= 0.058), the TBPI group (F_1.869, 11.21_ = 2.081, p= 0.172), the TBPI-I group (F_1.444, 2.888_ = 2.405, p= 0.234), or the TBPI-UI (F_1.441, 4.324_ = 1.249, p= 0.351). However, the post-hoc Dunnett’s multiple comparisons test found significant inhibition at 100 ms (p = 0.004, 95% C.I. = [11.07; 51.13]) for the CG. In the intergroup comparison, two-way ANOVA showed a main effect for Group when comparing CG x TBPI (F_1, 64_ = 4.112, p= 0.047), CG x TBPI-UI (F_1, 52_ = 7.361, p= 0.009) and CG x TBPI-I x TBPI-UI (F_2, 60_ = 4.230, p= 0.019) but not when comparing CG x TBPI-I (F_1, 48_ = 0.02186, p= 0.883) or TBPI-I x TBPI-UI (F_1, 20_= 3.168, p= 0.090). Post-hoc tests revealed no significant pairwise differences between different groups for each ISI.

Figure 3 shows means and standard errors for normalized MEP amplitudes in respect of SAI (Figure 3-A) and LAI (Figure 3-B) intervals after peripheral electrical stimulation applied to the face for each group. For the CG, inhibition was observed at 15, 35, 45, 55 and 65 ms, with maximum inhibition at 65 ms (21.19% ± 6.67). The TBPI group displayed slight inhibition at 55 ms (11.67% ± 10.76) and a slight facilitation at 15 ms (9.8% ± 7.49). After group separation, the TBPI-I group showed facilitation at 15 ms (13.40% ± 3.99) and the TBPI-UI group, at 25 ms (7.90% ± 5.05). RM one-way ANOVA for the face-hand interaction showed no significant difference within the groups (CG: F_2.418, 24.18_ = 1.268, p= 0.304; TBPI: F_2.496, 14.98_ = 1.229, p= 0.329; TBPI-I: F_1.113, 2.227_ = 1.342, p= 0.366; TBPI-UI: F_1.458, 4.374_ = 0.6063, p= 0.537). But the post-hoc Dunnett’s multiple comparisons test found significant inhibition at 65 ms (p = 0.042, 95% C.I. = [0.7010; 41.69]) for the CG. Two way ANOVA also showed a main effect for Group when comparing CG x TBPI (F_1, 96_ = 4.179, p= 0.044) and CG x TBPI-UI (F_1, 78_ = 5.665, p= 0.020) but not when comparing CG x TBPI-I (F_1, 72_ = 0.5113, p= 0.477), TBPI-I x TBPI-UI (F_1, 30_ = 1.209, p= 0.280) or the three groups (CG x TBPI-I x TBPI-UI: F_2, 90_ = 2.698, p= 0.073). Once again, post-hoc tests revealed no significant pairwise differences between different groups for each ISI.

For face-hand LAI intervals, there was no inhibition or facilitation for the CG, but a facilitation effect was observed for the TBPI group at all ISI, with a maximum facilitation of 39.90% ± 15.40 at 100 ms. TBPI-I and TBPI-UI groups, once subdivided, also displayed facilitation (55.20% ± 24.74 at 100 ms; 38.30% ± 32.55 at 200 ms; 10.90% ± 7.16 at 400 ms and 28.40% ± 20.53 at 100 ms; 9.5% ± 4.91 at 300 ms; 35.00% ± 12.80 at 400 ms, respectively). RM one-way ANOVA showed no significant difference within the groups for LAI intervals (CG: F_2.871, 28.71_ = 0.2152, p= 0.878; TBPI: F_1.815, 10.89_ = 3.184, p= 0.085; TBPI-I: F_1.402, 2.804_ = 2.552, p= 0.225; TBPI-UI: F_1.691, 5.073_ = 2.381, p= 0.187). For intergroups comparisons there was a significant effect for Group between CG x TBPI (F_1, 64_ = 12.34, p= 0,0008), CG x TBPI-I (F_1, 48_ = 9.068, p= 0.004), CG x TBPI-UI (F_1, 52_ = 6.887, p= 0.011) and CG x TBPI-I x TBPI-UI (F_2, 60_ = 6.381, p= 0.003) but not when comparing TBPI-I x TBPI-UI (F_1, 20_ = 0.4434, p= 0.513). Post-hoc tests revealed significant pairwise differences when comparing the 100 ms ISI between CG x TBPI (p = 0.015, 95% C.I. = [-82,49; -6.508]), CG x TBPI-I (p = 0.021, 95% C.I. = [-112.7; -6.890]) and CG x TBPI-I x TBPI-UI (p = 0.011, 95% C.I. = [-108.0; -11.68] for the CG x TBPI-I comparison).

Table S.2 in the Supplementary Material shows the mean and standard error MEP peak-to-peak amplitudes for all conditions and experimental blocks for each group. Figure S.2 in the Supplementary Material shows individual normalized mean MEP amplitudes for each participant within all groups for all experimental blocks.

### 3.4 Afferent facilitation in face-hand sensorimotor circuits correlate with pain and functional characteristics in TBPI patients

In order to perform correlation analysis, intervals with the greatest modulatory effect for the TBPI group were selected for each block: 45 ms for hand SAI, 100 ms for hand LAI, 65 ms for face-to-hand SAI and 100 ms for face-to-hand LAI. Pearson correlation coefficients were obtained to assess if there was a linear relationship with clinical data, questionnaire results and pain scores. Among the TBPI group, normalized mean MEP amplitudes at the 100 ms ISI for the face-to-hand interaction were positively correlated with CSI (r = 0.643, p = 0.031) and DASH (r = 0.614, p = 0.039) scores (Figure 4-A and 4-B).

**Figure 4.**
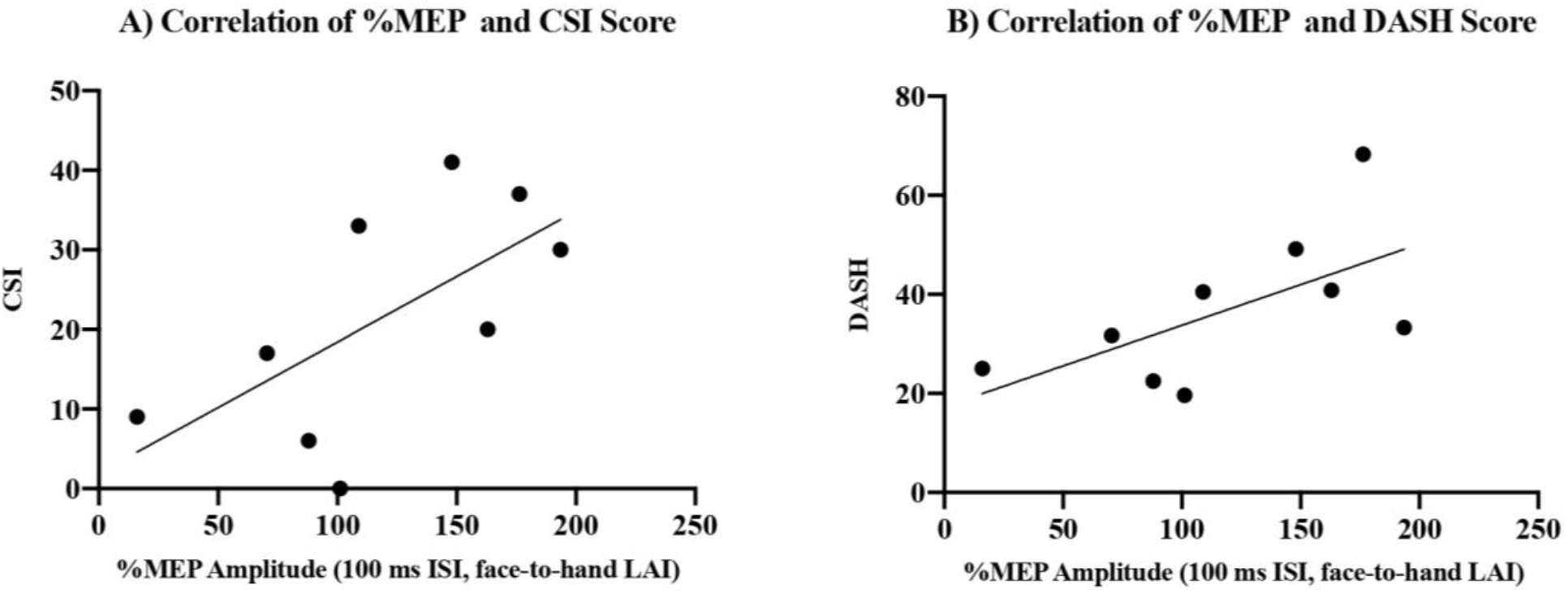
Correlation analysis. The interstimulus intervals with the greatest modulatory effect for the TBPI group were selected for each block: 45 ms for hand SAI, 100 ms for hand LAI, 65 ms for face-to-hand SAI and 100 ms for face-to-hand LAI. Individual normalized mean amplitudes values for the selected ISI were used to assess if there was a linear relationship with clinical data (age, time between injury and surgery, time between surgery and participation on the experiment and scores in the EHI, MMSE, DASH, PCL-C, VAS, DN4, BPI, CSI and McGill questionnaires). Pearson correlation coefficients were obtained. For the face-hand interaction, at long latency intervals, there was a positive correlation between %facilitation at the 100 ms ISI and the CSI (r = 0.643, p = 0.031) and DASH (r = 0.614, p = 0.039) scores.

## 4 Discussion

The AI paradigm was applied for the first time in patients with TBPI, a severe PNS injury that leads to upper limb sensorimotor impairment, chronic pain and plastic changes in cortical representations. To assess sensorimotor integration in the hand, a peripheral electrical stimulation was applied to the tip of the index finger followed by TMS at the contralateral FDI hot spot. In addition, sensorimotor integration between the face and the hand was assessed through peripheral electrical stimulation applied to the face followed by the TMS pulse over the contralateral FDI hot spot. We observed preserved hand sensorimotor integration for TBPI patients at short latency intervals (SAI), but not at long latency intervals (LAI), especially not in the TBPI-UI group. In respect of face-to-hand sensorimotor integration, the results showed no inhibition at SAI intervals for the TBPI patients. Once again, this was more evident for the TBPI-UI group. Lastly, for face-to-hand long latency intervals, a facilitation effect was observed in the TBPI patients. This long afferent facilitation was more prominent for the TBPI-I group and positively correlated with CSI and DASH scores.

### 4.1 Hand sensorimotor integration and SAI in controls and in TBPI patients

In our study, we verified typical sensorimotor integration through the observation in the control group (CG) of a significant SAI in the hand at different ISI (15, 25, 35, 45, and 55 ms). Our findings agreed with those of other studies that reported that the electrical stimulation of a digital nerve in the index finger evokes an SAI from 20 to 50 ms in the FDI.” (Tamburin et al., 2002; Tamburin et al., 2005). SAI reflects the interaction between the somatosensory and the motor systems, which is marked by the importance of sensory feedback for motor control (Scott et al., 2015). Previous studies using tractography techniques have shown that U-shaped fronto-parietal tracts connect the pre and postcentral gyrus in homologous representation areas of the primary somatosensory cortex (S1) and the primary motor cortex (M1), thus providing the anatomical substrate for the exchange of information between the somatosensory and the motor cortices (Catani et al., 2012). Other routes between the two regions exist, as signals detected by cutaneous and proprioception receptors are transmitted to contralateral thalamic nuclei and from there on to the M1 (Ruddy et al., 2016). It is possible that the sensorimotor integration modulatory effect known as SAI happens through the neuronal projections between the S1 and the M1, recruiting M1 interneurons capable of inhibiting pyramidal neurons. Another possibility is that SAI results from the activation of the direct projections between the sensory thalamus and the M1 (Hari et al., 1984; Nagamatsu et al., 2001; Turco et al., 2018).

SAI has been used as a tool to investigate changes in sensorimotor functions in different pathologies and injuries. The relationship between SAI reduction and conditions with impaired cognitive functions characterized by cholinergic dysfunction, such as Parkinson’s, Alzheimer’s and other types of dementia or neurological disorders such as ischemic or hemorrhagic stroke, traumatic brain injury, spinal cord injury, and schizophrenia has already been well documented (Di Lazzaro et al., 2000; Di Lazzaro et al., 2002; Di Lazzaro et al., 2004; Di Lazzaro et al., 2005; Sakuma, Murakami, and Nakashima, 2007; Nardone et al., 2008; Celebi et al., 2012). However, the TBPI patients in our study displayed significant SAI at the hand. In addition, there was no significant difference in respect of hand SAI when comparing the TBPI groups and the CG, which might indicate that TBPI patients, even those assessed on their injured limb (TBPI-I group), somehow have preserved sensorimotor integration for the hand, despite their peripheral neurological injury. Intracortical circuit dynamics offer one potential explanation for this phenomenon, with Alle et al. (2009) demonstrating an interaction between SAI and intracortical inhibitory mechanisms. Short intracortical inhibition (SICI) and SAI were examined by measuring MEP inhibition at a hand muscle after a sub-threshold TMS pulse or after a peripheral electrical stimulation at the ulnar nerve, respectively. Each of the stimuli applied alone had similar effects, but SICI was reduced or even disinhibited when the peripheral electrical stimulation was co-applied, and SAI was reduced or disinhibited when the sub-threshold TMS pulse was co-applied. The authors suggested that SICI and SAI are, therefore, mediated through reciprocally connected GABAergic inhibitory interneurons (Alle et al., 2009). Moreover, Cash et al. (2015) investigated the influence of SAI on excitatory interneuronal cortical circuits and found that short-interval intracortical facilitation (SICF) was facilitated in the presence of SAI, suggesting that there is also a relationship between SAI and SICF. Thus, in situations in which intracortical disinhibition is present, there could be a decrease in the inhibitory control that SICI exerts over SAI, resulting in SAI exacerbation. In fact, it has been demonstrated that the transection of peripheral nerves can produce a dramatic reduction in GABA in S1 resulting in the loss of GABAergic intracortical inhibition (Garraghty, LaChica, and Kaas, 1991).

Another interesting finding was that TBPI patients, especially those in the TBPI-I group, displayed hand SAI at longer ISI (45 and 55 ms) when compared to the CG. Most hand SAI studies have reported the occurrence of inhibition at intervals that correspond to the arrival of the afferent stimulus at the S1 (Tokimura et al., 2000; Bikmullina et al., 2009; Asmussen et al., 2013). A possible explanation for our finding is that TBPI patients have altered nerve conduction velocity due to the injured pathway (Ferrante and Wilbourn, 2002), causing a delay in the arrival of afferent information at the S1 so that only longer ISI were able to match the motor output. In this sense, a useful approach would be to pair the peripheral electrical stimulation with EEG recordings, so that ISI choice could be based on the latency of the N20 component of the somatosensory evoked potential (Di Lazzaro et al., 2002; Di Lazzaro et al., 2005; Alle et al., 2009; Ferreri et al., 2012; Cash et al., 2015).

### 4.2 Hand sensorimotor integration and the TBPI effect on LAI

Hand sensorimotor integration can also be assessed at longer intervals through LAI. In our study, LAI was present for the CG group at 100 ms, a shorter interval than that described for the nerve stimulation at the middle finger, which is capable of generating LAI at the FDI at intervals ranging from 200 to 600 ms (Chen, Corwell, and Hallett, 1999; Turco et al., 2018). However, hand LAI was not observed for the TBPI group. The comparisons between groups pointed to a significant difference between the CG and the TBPI group and between the CG and the TBPI-UI group, which displayed a tendency towards facilitation at longer ISI (300 ms). LAI circuits involve generalized activation of several cortical areas beyond the S1, such as the posterior parietal cortex and the secondary somatosensory cortex. These areas project to the M1, where they mediate MEP inhibition, but neural pathways between basal nuclei, thalamus, and the cortex may also be involved in LAI (Chen, Corwell, and Hallett, 1999; Turco et al., 2018). Therefore, the absence of LAI in the TBPI patients is in line with previous studies that indicated that the connectivity between sensorimotor cortices and higher-order areas seem to be compromised in TBPI (Qiu et al., 2014; Lu et al., 2016; Yechen Lu et al., 2016; Bhat et al., 2017; Lyu et al., 2017).

Afferent facilitation has been less explored in literature. Studies suggest that an excitatory effect could occur at intervals from 25 to 80 ms for hand muscles after the electrical stimulation of the median nerve, but not after the electrical stimulation of the index finger (Deletis et al., 1992; Devanne et al. 2009; Deveci et al., 2020). In our study, the facilitation tendency observed for the TBPI-UI group was elicited with much longer ISI, so it probably does not reflect activity in these previously described afferent facilitation circuits but might be due to the GABA reduction caused by the nerve injury, resulting in a hyperexcitability state that led to the observed facilitation. TBPI-UI patients seemed more affected by this than TBPI-I patients. It is worth noting that the TBPI-UI group was, by experimental design, the group with more severe injuries (i.e., complete TBPI). Thus, even though they were evaluated on their uninjured limb, bilateral cortical disinhibition beyond the S1 caused by the peripheral injury could be the cause of the facilitatory effect observed.

### 4.3 Face-to-hand sensorimotor integration and the TBPI effect on SAI and LAI

Although less explored, afferent inhibition has also been previously reported in face muscles following the peripheral electrical stimulation of the trigeminal nerve (face-to-face SAI) but not after the stimulation of the facial nerve (Pilurzi et al., 2020). Furthermore, face-to-hand SAI was also observed for healthy subjects, although the LAI effect was not investigated (Ramalho et al., 2022). The CG in our study displayed face-to-hand inhibition at the 65 ms ISI, but not at LAI intervals. For the TBPI groups, face-to-hand SAI was shown to be missing. It was especially absent for the TBPI-UI group. Just as for the CG, face-to-hand interaction at long latency intervals resulted in no inhibition for the TBPI groups. On the contrary, there was a marked tendency towards facilitation, as revealed by a significant facilitation at the 100 ms ISI in the comparison between the CG and the TBPI group and between the CG and the TBPI-I group, a process we termed long-latency afferent facilitation (LAF). Again, it should be emphasized that bilateral changes in cortical representations have been reported for TBPI (Hsieh et al., 2002; Yoshikawa et al., 2012; Hua et al., 2013; Liu et al., 2013; Qiu et al., 2014; Fraiman et al., 2016; Kakinoki et al., 2017). Therefore, bilateral cortical hyperexcitability resulting from TBPI could explain the absence of face-to-hand SAI and the presence of face-to-hand LAF in both TBPI groups.

Altered face-to-hand AI in TBPI patients might relate to a mechanism proposed in some studies through which the changes in functional boundaries that follow peripheral lesions results in sensorimotor cortex reorganization (i.e. the unmasking of pre-existing, normally inhibited, connections between different body parts) (Jacobs and Donoghue, 1991; Ramachandran et al., 1993; Farnè et al., 2002; Li et al., 2014; Harding-Forrester and Feldman, 2018). Interestingly, amputees with the ability to experience phantom movement have preserved M1 representations of muscles of the absent limb (Roux et al., 2003; Mercier et al., 2006; Raffin et al., 2016). Moreover, stump muscle activity recorded during phantom hand movements suggests that the amputated hand cortical representation overlaps with the cortical representation of the stump (Reilly et al., 2006; Reilly and Sirigu, 2008), indicating a massive reorganization of the body representation in the brain after a peripheral injury. In fact, the persistence of cortical representations of the hand after an amputation may be related to the ability to functionally recover and return to the original representations after hand allograft (Vargas et al., 2009). Although recent research on arm amputation failed in identifying a clear sensorimotor cortex remapping of lip representation into the missing hand territory during lip movements (Makin et al., 2013; Kikkert et al., 2018), in our study, bilateral cortical hyperexcitability resulting from TBPI could explain the absence of face-to-hand SAI and the presence of face-to-hand LAF in both TBPI groups. Moreover, it corroborates with the notion that multiple factors seem to interact to maintain local functional representations and long-range connectivity (Makin et al., 2013; Makin and Flor, 2020).

### 4.4 Effects of TBPI and pain over face-to-hand sensorimotor integration

Persistent hand representation and maladaptive cortical reorganization have both proven to be relevant to phantom limb pain (Flor et al., 1995; Karl et al., 2001), and altered sensorimotor integration might be a reflection of such mechanisms. Phantom limb pain, originally described in amputees, is also present in cases of nerve avulsion, with approximately 40% of TBPI caused by avulsion present with phantom limb pain (Parry, Wynn, and Wynn Parry, 1980; Melzack, 1992; Ramachandran and Hirstein, 1998; Shankar, Hansen, and Thomas, 2015; Teixeira et al., 2015). Neuropathic pain, present in phantom limb pain but not restricted to it, is the most frequently described pain mechanism in TBPI, with an incidence of around 70% according to Narakas (1985). It can develop immediately after the injury, which suggests a relationship with deafferentation, or evolve over months and worsening over time, which indicates the involvement of nervous system plasticity mechanisms (Lovaglio et al., 2019). Moreover, chronic neuropathic pain has a distinct role in cortical plasticity and in the inhibition/disinhibition circuits balance (Schwenkreis et al., 2010; Gustin et al., 2012; Caumo et al., 2016; Parker et al., 2016).

Some chronic pain conditions have been examined with AI protocols with divergent results. There was no difference in AI for patients with focal hand dystonia (Avanzino et al., 2008) or complex regional pain syndrome (Morgante et al., 2017), but there was a decrease in AI in patients with cervical dystonia (Zittel et al., 2015), and chronic shoulder pain (Bradnam et al., 2016). In an experimental pain scenario, SAI was similar for both painful and non-painful conditions (Mercier et al., 2016). Considering these results, it is possible that neuropathic pain and pain from other origins might influence AI differently. For the TBPI patients in our study, there was a positive correlation between CSI and DASH scores and normalized mean MEP amplitudes at 100 ms in the face-to-hand interaction. The CSI inventory is a health symptoms questionnaire developed to screen patients at risk of central pain sensitization with scores above 40 points indicating the presence of central sensitization (Caumo et al., 2017). The DASH questionnaire assesses upper limb functionality and its score ranges from 0 (maximum functionality) to 100 (maximum disability) (Orfale et al., 2005). Our results indicate that face-to-hand LAF may, therefore, reflect altered functionality and the presence of central sensitization pain, which may also influence each other. Central sensitization is one of the most important mechanisms for chronic pain, including neuropathic pain, and studies have indicated that neuropathic pain is accompanied by a reduced GABAergic inhibitory function (Caijuan et al., 2019), which was reflected in the hyperexcitability of the sensorimotor face-to-hand circuits we observed. Finally, as mentioned above, face-to-hand LAF may also reflect maladaptive cortical reorganization, one of the proposed mechanisms for the origins of phantom limb pain, a phenomenon present in nerve avulsions events such as complete TBPI.

## 5 Limitations

The main limitation of the study was the small number and the heterogeneity of the TBPI participants. However, the decision not to enroll more TBPI participants after the start of the COVID-19 pandemic was made to ensure everyone’s safety. This limitation made conclusions about AI in the population with TBPI as a whole more difficult. Moreover, in future studies it might be useful to restrict the clinical characteristics of the TBPI patients enrolled to reduce the variability generated by participants’ heterogeneity.

## 6 Conclusions

This study presents the first evidence in respect of how TBPI and its consequences influence afferent inhibition - the interaction between somatosensory and motor systems in the hand and between the face and the hand. Preserved SAI in the hand for TBPI patients was observed, a sign that the peripheral injury did not prevent sensorimotor integration. Hand LAI, however, was reduced for TBPI patients, especially for those with complete injury who were assessed in their uninjured upper limb, suggesting that higher-order regions involved in sensorimotor integration are affected by the peripheral injury.

Our results showed bilateral central reorganization involving hand and face sensorimotor representations. This was observed through decreased face-to-hand SAI for the TBPI groups, which is an indication that plastic changes resulting from the nerve injury had a direct influence over face-to-hand sensorimotor circuits. For long latency intervals, a facilitation effect was observed for both TBPI groups which positively correlated with the degree of functionality and the presence of central sensitization pain. These results point to the existence of an inhibitory regulation system between the representations of the face and the hand that seems to be suppressed in TBPI.

These findings also reinforce the idea that changes arising from TBPI are not restricted to the injured limb and that cortical alterations resulting from a unilateral peripheral injury can extend to both brain hemispheres.

## Data Availability

All data produced in the present study are available upon reasonable request to the authors

## 8 Author Contributions

F.F.T., B.L.R., K.T.R. and C.D.V. conceived and planned the experiments. F.F.T., M.R.R., A.C.S. and V.H.F.M.F. carried out the experiments. F.F.T., M.R.R., A.C.S. and V.H.F.M.F. contributed to data processing. F.F.T. performed data analysis. F.F.T., B.L.R., R.P.C. and C.D.V. contributed to the interpretation of the results. F.F.T. took the lead in writing the manuscript. All authors provided critical feedback and helped shape the research, analysis and manuscript.

