## Supplemental Material captions for "Plasticity of the face-hand sensorimotor circuits after a traumatic brachial plexus injury"

**Supplementary Material**

**Table S.1.**

Lost trials in each block for every participant, according to the established criteria (see text for details). Notation in each cell displayed as: total of lost trials (the condition in which the loss occurred) or total of lost trials (condition: number of lost events in that condition), being 1= control condition (transcranial magnetic stimulation without previous peripheral electrical stimulation), 2 = 15 ms for SAI and 100 ms for LAI, 3 = 25 ms for SAI and 200 ms for LAI, 4 = 35 ms for SAI and 300 ms for LAI, 5 = 45 ms for SAI and 400 ms for LAI, 6 = 55 ms, and 7 = 65 ms. *participant excluded due to excess losses, **participant excluded for not meeting MEP > 300mV criterion, ***participant excluded in the temporal analysis.

**Table S.2.**

Results presented as mean ± standard error. ISI, interstimulus interval; CTRL, control condition (transcranial magnetic stimulation without previous peripheral electrical stimulation); TBPI group, all traumatic brachial plexus injury participants; TBPI-I group, TBPI patients assessed on the injured side; TBPI-UI, TBPI patients assessed on the uninjured side.

**Figure S.1.**

Data processing details. Left column displays all trials after removing those with evidence of muscle contraction or electromyography artifacts, presented for all interstimulus intervals and the control condition for each participant in all groups: A- Hand SAI, B- Hand LAI, C- Face-hand SAI, D- Face-hand LAI. Center column displays all remaining trials after outliers removal: E- Hand SAI, F-Hand LAI, G- Face-hand SAI, H- Face-hand LAI. Right column displays data after exclusion of participants in each block: I- Hand SAI, J- Hand LAI, K- Face-hand SAI, L- Face-hand LAI. Circles represent Control Group participants, triangles represent TBPI-I group participants (traumatic brachial plexus injury participants assessed on the injured side) and squares represent TBPI-UI group participants (traumatic brachial plexus injury participants assessed on the uninjured side). Details presented in Materials and Methods section in the text.

**Figure S.2**

Results presented by group with individual mean normalized values. Left column displays results for the Control Group for hand SAI (A), hand LAI (B), face-hand SAI (C) and face-hand LAI (D). Center column displays results for the TBPI-I group (traumatic brachial plexus injury patients assessed on the injured side) for hand SAI (E), hand LAI (F), face-hand SAI (G) and face-hand LAI (H). Right column displays results for the TBPI-UI group (traumatic brachial plexus injury patients assessed on the uninjured side). Values below the dotted line at 100% indicate an inhibition effect, values above the dotted line indicate a facilitation effect.
