## Supplemental Table 1 for "Plasticity of the face-hand sensorimotor circuits after a traumatic brachial plexus injury"

**Table S1. Exclusion Process Summary**

| Participant | Experiment | Mean MEP Control (mV) | Excluded - Muscle Contraction | Missing Trials | Excluded - EMG Artifact | Excluded - Outlier | Total Lost Trials | Decision |
|---|---|---|---|---|---|---|---|---|
| TBPI-I1 | 1 | 0.837 | 0 | 2 (5; 6) | 0 | 6 (3:1; 6:3, 7:2) | 8 (3:1; 5:1; 6:4; 7:2) | Sustained |
| | 2 | 0.929 | 0 | 0 | 0 | 3 (2:1; 3:1; 5:1) | 3 (2:1; 3:1; 5:1) | Sustained |
| | 3 | 0.789 | 1 (2) | 0 | 0 | 3 (3:1; 4:2) | 4 (2:1; 3:1; 4:2) | Sustained |
| | 4 | 0.428 | 0 | 0 | 0 | 3 (1:1; 5:2) | 3 (1:1; 5:2) | Sustained |
| TBPI-I2 | 1 | 1.048 | 0 | 0 | 6 (2:6) | 9 (4:3; 5:2; 6:2; 7:2) | 15 (2:6; 4:3; 5:2; 6:2; 7:2) | Sustained |
| | 2 | 0.628 | 0 | 1 (4) | 44 (1:7; 2:10; 3:14; 4:3; 5:10) | 5 (2:1; 4:4) | 49 (1:7; 2:11; 3:14; 4:7; 5:10) | Excluded* |
| | 3 | 0.943 | 51 (1:6; 2:6; 3:5; 4:8; 5:10; 6:6; 7:10) | 0 | 0 | 2 (3:2) | 53 (1:6; 2:6; 3:7; 4:8; 5:10; 6:6; 7:10) | Excluded* |
| | 4 | 0.889 | 54 (1:11; 2:12; 3:12; 4:8; 5:11) | 0 | 0 | 1 (4) | 55 (1:11; 2:12; 3:12; 4:9; 5:11) | Excluded* |
| TBPI-I3 | 1 | 1.073 | 6 (1:2; 3:2; 4:1; 7:1) | 0 | 0 | 0 | 6 (1:2; 3:2; 4:1; 7:1) | Sustained |
| | 2 | 1.054 | 1 (5) | 0 | 0 | 1 (1) | 2 (1:1;5:1) | Sustained |
| | 3 | 0.948 | 1 (4) | 0 | 0 | 2 (1:1; 4:1) | 3 (1:2; 4:1) | Sustained |
| | 4 | 1.204 | 6 (3:3; 4:2; 5:1) | 0 | 0 | 2 (4:1; 5:1) | 8 (3:3; 4:3; 5:2) | Sustained |
| TBPI-I4 | 1 | 0.221 | 0 | 1 (5) | 14 (2:5; 3:8; 4:1) | 13 (1:1; 2:1; 4:3; 5:3; 6:2; 7:3) | 28 (1:1; 2:6; 3:8; 4:3; 5:4; 6:2; 7:3) | Excluded** |
| | 2 | 0.125 | 0 | 0 | 0 | 7 (1:2; 2:1; 3:1; 4:1; 5:2) | 7 (1:2; 2:1; 3:1; 4:1; 5:2) | Excluded** |
| | 3 | 0.144 | 0 | 0 | 0 | 7 (2:1; 3:1; 5:2; 6:3) | 7 (2:1; 3:1; 5:2; 6:3) | Excluded** |
| | 4 | 0.128 | 0 | 0 | 0 | 5 (1:1; 2:2; 3:1; 5:1) | 5 (1:1; 2:2; 3:1; 5:1) | Excluded** |
| TBPI-I5 | 1 | 0.735 | 0 | 0 | 0 | 3 (2:1; 3:1; 6:1) | 3 (2:1; 3:1; 6:1) | Sustained |
| | 2 | 0.717 | 0 | 0 | 0 | 6 (1:1; 2:2; 4:1; 5:2) | 6 (1:1; 2:2; 4:1; 5:2) | Sustained |
| | 3 | 0.971 | 1 (5) | 0 | 0 | 7 (1:1; 2:2; 3:1; 4:3) | 8 (1:1; 2:2; 3:1; 4:3; 5:1) | Sustained |
| | 4 | 0.795 | 3 (2:2; 3:1) | 0 | 0 | 4 (1:2; 2:2) | 7 (1:2; 2:4; 3:1) | Sustained |
| TBPI-UI1 | 1 | 0.961 | 0 | 1 (4) | 0 | 4 (2:1; 4:1; 5:1; 6:1) | 5 (2:1; 4:2; 5:1; 6:1) | Sustained |
| | 2 | 1.876 | 0 | 0 | 0 | 0 | 0 | Sustained |
| | 3 | 1.738 | 0 | 0 | 0 | 3 (4:1; 5:2; 6:1) | 3 (4:1; 5:2; 6:1) | Sustained |
| | 4 | 1.710 | 0 | 0 | 0 | 0 | 0 | Sustained |
| TBPI-UI2 | 1 | 0.797 | 0 | 0 | 0 | 8 (1:1; 2:1; 3:3; 4:1; 5:1; 6:1) | 8 (1:1; 2:1; 3:3; 4:1; 5:1; 6:1) | Sustained |
| | 2 | 0.353 | 2 (1:2) | 0 | 0 | 4 (1:2; 2:1; 3:1) | 6 (1:4; 2:1; 3:1) | Sustained |
| | 3 | 1.626 | 0 | 0 | 0 | 10 (1:2; 2:2; 3:2; 4:1; 7:3) | 10 (1:2; 2:2; 3:2; 4:1; 7:3) | Sustained |
| | 4 | 1.626 | 0 | 0 | 0 | 4 (1:1; 2:1; 4:2) | 4 (1:1; 2:1; 4:2) | Sustained |
| TBPI-UI3 | 1 | 0.699 | 0 | 0 | 12 (1:2; 2:1; 4:3; 5:5; 6:1) | 7 (3:2; 5:1; 6:2; 7:2) | 29 (1:2; 2:1; 3:2; 4:3; 5:6; 6:3; 7:2) | Sustained |
| | 2 | 0.617 | 0 | 0 | 10 (2:4; 3:3; 4:3) | 4 (3:1; 4:1; 5:2) | 14 (2:4; 3:4; 4:4; 5:2) | Sustained |
| | 3 | 0.689 | 1 (3) | 0 | 0 | 3 (1:1; 3:1; 7:1) | 4 (1:1; 3:2; 7:1) | Sustained |
| | 4 | 1.052 | 2 (5:2) | 0 | 0 | 5 (1:1; 2:1; 3:1; 4:2) | 7 (1:1; 2:1; 3:1; 4:2; 5:2) | Sustained |

| | | | | | | | | |
|---|---|---|---|---|---|---|---|---|
| TBPI-UI4 | 1 | 0.577 | 0 | 0 | 0 | 4 (3:1; 4:1; 6:1; 7:1) | 4 (3:1; 4:1; 6:1; 7:1) | Sustained |
| | 2 | 0.459 | 0 | 0 | 0 | 7 (1:2; 2:1; 3:2; 5:2) | 7 (1:2; 2:1; 3:2; 5:2) | Sustained |
| | 3 | 0.501 | 0 | 0 | 0 | 9 (1:2; 2:1; 3:1; 4:2; 5:1; 6:1; 7:1) | 9 (1:2; 2:1; 3:1; 4:2; 5:1; 6:1; 7:1) | Sustained |
| | 4 | 0.415 | 1 (3) | 0 | 0 | 5 (1:2; 3:1; 4:2) | 6 (1:2; 3:2; 4:2) | Sustained |
| Control 1 | 1 | 0.943 | 0 | 1 (5) | 0 | 0 | 1 (5:1) | Excluded*** |
| | 2 | 1.544 | 0 | 0 | 0 | 4 (1:3; 3:1) | 4 (1:3; 3:1) | Sustained |
| | 3 | 0.423 | 0 | 0 | 0 | 5 (1:1; 6:2; 7:2) | 5 (1:1; 6:2; 7:2) | Excluded*** |
| | 4 | 0.598 | 0 | 0 | 0 | 6 (4:3; 5:3) | 6 (4:3; 5:3) | Sustained |
| Control 2 | 1 | 0.794 | 0 | 0 | 0 | 7 (1:2; 2:1; 5:2; 7:2) | 7 (1:2; 2:1; 5:2; 7:2) | Excluded*** |
| | 2 | 1.327 | 0 | 0 | 0 | 1 (5) | 1 (5:1) | Sustained |
| | 3 | 1.439 | 22 (1:3; 2:1; 3:2; 4:4; 5:4; 6:5; 7:3) | 0 | 0 | 0 | 22 (1:3; 2:1; 3:2; 4:4; 5:4; 6:5; 7:3) | Sustained |
| | 4 | 1.497 | 1 (4) | 1 (5) | 0 | 2 (1:2) | 4 (1:2; 4:1; 5:1) | Sustained |
| Control 3 | 1 | 1.467 | 3 (1:1; 2:1; 7:1) | 0 | 0 | 5 (1:4; 3:1) | 8 (1:5; 2:1; 3:1; 7:1) | Sustained |
| | 2 | 1.814 | 8 (1:2; 2:1; 3:1; 4:2; 5:2) | 0 | 0 | 1 (2) | 9 (1:2; 2:2; 3:1; 4:2; 5:2) | Sustained |
| Control 4 | 1 | 0.171 | 2 (3:1; 5:1) | 1 (6) | 0 | 10 (1:2; 2:2; 4:1; 5:1; 6:3; 7:1) | 12 (1:2; 2:2; 3:1; 4:1; 5:2; 6:3; 7:1) | Excluded** |
| | 2 | 0.129 | 0 | 0 | 0 | 4 (1:1; 3:2; 5:1) | 4 (1:1; 3:2; 5:1) | Excluded** |
| | 3 | 0.175 | 0 | 0 | 0 | 14 (1:5; 2:2; 4:2; 5:1; 6:3; 7:1) | 14 (1:5; 2:2; 4:2; 5:1; 6:3; 7:1) | Excluded** |
| | 4 | 0.131 | 1 (4) | 0 | 0 | 4 (1:2; 3:1; 5:1) | 5 (1:2; 3:1; 4:1; 5:1) | Excluded** |
| Control 5 | 1 | 0.105 | 0 | 0 | 0 | 11 (1:2; 2:2; 3:2; 4:1; 5:2; 6:1; 7:1) | 11 (1:2; 2:2; 3:2; 4:1; 5:2; 6:1; 7:1) | Excluded** |
| | 2 | 0.174 | 0 | 0 | 0 | 7 (1:1; 2:1; 3:2; 4:1; 5:2) | 7 (1:1; 2:1; 3:2; 4:1; 5:2) | Excluded** |
| Control 6 | 1 | 0.049 | 0 | 0 | 0 | 7 (1:3; 2:1; 4:2; 5:1) | 7 (1:3; 2:1; 4:2; 5:1) | Excluded** |
| | 2 | 0.249 | 0 | 0 | 0 | 1 (1) | 1 (1) | Excluded** |
| | 3 | 0.385 | 0 | 0 | 0 | 5 (2:1; 3:2; 5:1; 7:1) | 5 (2:1; 3:2; 5:1; 7:1) | Sustained |
| | 4 | 0.163 | 0 | 0 | 0 | 4 (1:1; 2:2; 5:1) | 4 (1:1; 2:2; 5:1) | Excluded** |
| Control 7 | 1 | 0.617 | 0 | 0 | 3 (2:1; 3:2) | 7 (1:1; 2:1; 4:1; 5:3; 6:1) | 10 (1:1; 2:2; 3:2; 4:1; 5:3; 6:1) | Sustained |
| | 2 | 1.127 | 0 | 0 | 0 | 1 (3) | 1 (3) | Sustained |
| | 3 | 1.167 | 0 | 0 | 0 | 0 | 0 | Sustained |
| | 4 | 1.279 | 0 | 0 | 0 | 3 (5:3) | 3 (5:3) | Sustained |
| Control 8 | 1 | 0.871 | 0 | 0 | 0 | 6 (1:1; 2:4; 4:1) | 6 (1:1; 2:4; 4:1) | Sustained |
| | 2 | 0.652 | 0 | 0 | 0 | 3 (2:2; 3:1) | 3 (2:2; 3:1) | Sustained |
| | 3 | 0.830 | 0 | 0 | 0 | 3 (3:1; 4:1; 6:1) | 3 (3:1; 4:1; 6:1) | Sustained |
| | 4 | 0.609 | 0 | 0 | 0 | 0 | 0 | Sustained |
| Control 9 | 1 | 0.324 | 0 | 0 | 0 | 3 (2:1; 4:1; 5:1) | 3 (2:1; 4:1; 5:1) | Sustained |
| | 2 | 0.516 | 0 | 0 | 0 | 5 (2:1; 5:4) | 5 (2:1; 5:4) | Sustained |

| | | | | | | | | |
|---|---|---|---|---|---|---|---|---|
| Control 9 | 3 | 1.238 | 0 | 0 | 0 | 7 (3:1; 4:2; 5:1; 6:1; 7:2) | 7 (3:1; 4:2; 5:1; 6:1; 7:2) | Sustained |
| | 4 | 0.850 | 0 | 0 | 0 | 7 (2:2; 3:1; 4:4) | 7 (2:2; 3:1; 4:4) | Sustained |
| Control 10 | 1 | 0.752 | 0 | 0 | 0 | 2 (3:2) | 2 (3:2) | Sustained |
| | 2 | 0.800 | 0 | 0 | 0 | 2 (3:2) | 2 (3:2) | Sustained |
| Control 11 | 1 | 0.606 | 0 | 0 | 0 | 5 (4:2; 5:1; 6:1; 7:1) | 5 (4:2; 5:1; 6:1; 7:1) | Sustained |
| | 2 | 0.119 | 0 | 0 | 0 | 4 (1:3; 3:1) | 4 (1:3; 3:1) | Excluded** |
| | 3 | 0.743 | 0 | 0 | 0 | 0 | 0 | Sustained |
| | 4 | 1.088 | 0 | 0 | 0 | 2 (4:1; 5:1) | 2 (4:1; 5:1) | Sustained |
| Control 12 | 1 | 0.999 | 0 | 0 | 0 | 4 (4:2; 5:2) | 4 (4:2; 5:2) | Sustained |
| | 2 | 1.064 | 5 (2:1; 3:2; 5:2) | 0 | 0 | 4 (1:1; 3:2; 5:1) | 9 (1:1; 2:1; 3:4; 5:3) | Sustained |
| | 3 | 0.404 | 4 (2:1; 3:1; 6:1; 7:1) | 0 | 0 | 4 (1:1; 2:1; 5:1; 6:1) | 8 (1:1; 2:2; 3:1; 5:1; 6:2; 7:1) | Sustained |
| | 4 | 0.369 | 0 | 0 | 0 | 0 | 0 | Sustained |
| Control 13 | 1 | 0.496 | 0 | 0 | 0 | 4 (2:1; 3:1; 6:1; 7:1) | 4 (2:1; 3:1; 6:1; 7:1) | Sustained |
| | 2 | 0.551 | 0 | 0 | 0 | 4 (2:3; 4:1) | 4 (2:3; 4:1) | Sustained |
| | 3 | 0.581 | 0 | 0 | 1 (1) | 10 (1:4; 3:1; 5:1; 6:2; 7:2) | 11 (1:5; 3:1; 5:1; 6:2; 7:2) | Sustained |
| | 4 | 1.452 | 0 | 0 | 0 | 1 (1) | 1 (1) | Sustained |
| Control 14 | 1 | 0.697 | 0 | 0 | 0 | 5 (2:2; 3:1; 6:2) | 5 (2:2; 3:1; 6:2) | Sustained |
| | 2 | 0.193 | 0 | 0 | 0 | 8 (1:3; 2:1; 3:1; 4:1; 5:2) | 8 (1:3; 2:1; 3:1; 4:1; 5:2) | Excluded** |
| | 3 | 0.415 | 0 | 0 | 0 | 6 (4:2; 6:2; 7:2) | 6 (4:2; 6:2; 7:2) | Sustained |
| | 4 | 0.646 | 0 | 0 | 0 | 4 (1:1; 2:1; 3:1; 4:1) | 4 (1:1; 2:1; 3:1; 4:1) | Sustained |
| Control 15 | 1 | 0.371 | 0 | 0 | 0 | 4 (2:1; 5:2; 6:1) | 4 (2:1; 5:2; 6:1) | Sustained |
| | 2 | 0.163 | 1 (1) | 0 | 0 | 5 (1:1; 2:1; 4:1; 5:2) | 6 (1:2; 2:1; 4:1; 5:2) | Excluded** |
| | 3 | 0.695 | 3 (2:1; 5:2) | 0 | 0 | 15 (1:2; 2:3; 3:1; 4:3; 5:2; 6:2; 7:2) | 18 (1:2; 2:4; 3:1; 4:3; 5:4; 6:2; 7:2) | Sustained |
| | 4 | 1.530 | 0 | 0 | 0 | 5 (2:1; 3:1; 4:2; 5:1) | 5 (2:1; 3:1; 4:2; 5:1) | Sustained |
| Control 16 | 1 | 1.259 | 0 | 0 | 0 | 7 (2:1; 4:1; 5:3; 6:1; 7:1) | 7 (2:1; 4:1; 5:3; 6:1; 7:1) | Sustained |
| | 2 | 1.015 | 0 | 0 | 0 | 6 (2:2; 4:1; 5:3) | 6 (2:2; 4:1; 5:3) | Sustained |
| Control 17 | 3 | 0.028 | 0 | 0 | 0 | 12 (1:2; 2:2; 3:2; 4:1; 5:2; 6:3) | 12 (1:2; 2:2; 3:2; 4:1; 5:2; 6:3) | Excluded** |
| | 4 | 0.218 | 1 (1) | 2 (1, 2) | 0 | 1 (2) | 4 (1:2; 2:2) | Excluded** |
| Control 18 | 1 | 1.348 | 6 (2:1; 3:2; 4:1; 6:1; 7:1) | 0 | 0 | 2 (2:1; 5:1) | 8 (2:2; 3:2; 4:1; 5:1; 6:1; 7:1) | Sustained |
| | 2 | 1.220 | 0 | 0 | 0 | 0 | 0 | Sustained |
| | 3 | 0.962 | 0 | 0 | 0 | 1 (4) | 1 (4:1) | Sustained |
| | 4 | 1.040 | 0 | 0 | 0 | 3 (2:1; 3:1; 5:1) | 3 (2:1; 3:1; 5:1) | Sustained |
