## Supplemental Table 2 for "Plasticity of the face-hand sensorimotor circuits after a traumatic brachial plexus injury"

| Table S2. Groups Mean Motor Evoked Potential Amplitude (mV) | | | | | |
|---|---|---|---|---|---|
| Experimental block | ISI | Control Group | TBPI Group | TBPI-I Group | TBPI-UI Group |
| Hand SAI | CTRL | 0.8172 ± 0.11 | 0.8410 ± 0.06 | 0.9233 ± 0.08 | 0.7586 ± 0.08 |
| | 15 | 0.5997 ± 0.10 | 0.8670 ± 0.09 | 0.9317 ± 0.15 | 0.8023 ± 0.11 |
| | 25 | 0.5942 ± 0.10 | 0.6219 ± 0.13 | 0.8258 ± 0.21 | 0.4179 ± 0.10 |
| | 35 | 0.5143 ± 0.09 | 0.5286 ± 0.13 | 0.6351 ± 0.25 | 0.4221 ± 0.08 |
| | 45 | 0.4748 ± 0. 09 | 0.4292 ± 0.11 | 0.4654 ± 0.22 | 0.3930 ± 0.09 |
| | 55 | 0.4759 ± 0.08 | 0.4529 ± 0.12 | 0.4308 ± 0.23 | 0.4749 ± 0.14 |
| | 65 | 0.6045 ± 0.08 | 0.5568 ± 0.13 | 0.5612 ± 0.25 | 0.5523 ± 0.13 |
| Hand LAI | CTRL | 1.057 ± 0.12 | 0.8578 ± 0.23 | 0.8998 ± 0.10 | 0.8263 ± 0.35 |
| | 100 | 0.7429 ± 0.12 | 0.7842 ± 0.23 | 0.6244 ± 0.19 | 0.9041 ± 0.39 |
| | 200 | 0.8435 ± 011 | 0.7886 ± 0.24 | 0.7446 ± 0.16 | 0.8217 ± 0.44 |
| | 300 | 0.9650 ± 0.14 | 0.9805 ± 0.24 | 0.9472 ± 0.22 | 1.005 ± 0.41 |
| | 400 | 1.006 ± 0.14 | 0.9907 ± 0.24 | 0.9556 ± 0.19 | 1.017 ± 0.42 |
| Face-to-hand SAI | CTRL | 0.8053 ± 0.11 | 1.037 ± 0.18 | 0.9027 ± 0.06 | 1.138 ± 0.32 |
| | 15 | 0.7140 ± 0.12 | 1.155 ± 0.23 | 1.027 ± 0.09 | 1.251 ± 0.41 |
| | 25 | 0.7677 ± 0.11 | 1.103 ± 0.22 | 0.8956 ± 0.11 | 1.259 ± 0.38 |
| | 35 | 0.7249 ± 0.10 | 0.9752 ± 0.23 | 0.8354 ± 0.19 | 1.080 ± 0.41 |
| | 45 | 0.6587 ± 0.10 | 1.098 ± 0.32 | 0.7564 ± 0.22 | 1.353 ± 0.53 |
| | 55 | 0.6719 ± 0.09 | 0.9344 ± 0.21 | 0.7257 ± 0.25 | 1.091 ± 0.32 |
| | 65 | 0.6399 ± 0.10 | 0.9697 ± 0.18 | 0.8374 ± 0.25 | 1.069 ± 0.27 |
| Face-to-hand LAI | CTRL | 0.9960 ± 0.12 | 1.033 ± 0.20 | 0.8088 ± 0.22 | 1.201 ± 0.30 |
| | 100 | 0.8984 ± 0.14 | 1.269 ± 0.14 | 1.145 ± 0.16 | 1.362 ± 0.22 |
| | 200 | 1.020 ± 0.15 | 1.133 ± 0.15 | 0.9826 ± 0.09 | 1.246 ± 0.25 |
| | 300 | 0.9264 ± 0.14 | 1.098 ± 0.19 | 0.8638 ± 0.17 | 1.274 ± 0.29 |
| | 400 | 0.9877 ± 0.14 | 1.251 ± 0.23 | 0.8653 ± 0.19 | 1.54 ± 0.33 |
