## Supplementary figures and images for "Plasticity of the face-hand sensorimotor circuits after a traumatic brachial plexus injury"

### Supplemental Figure 1

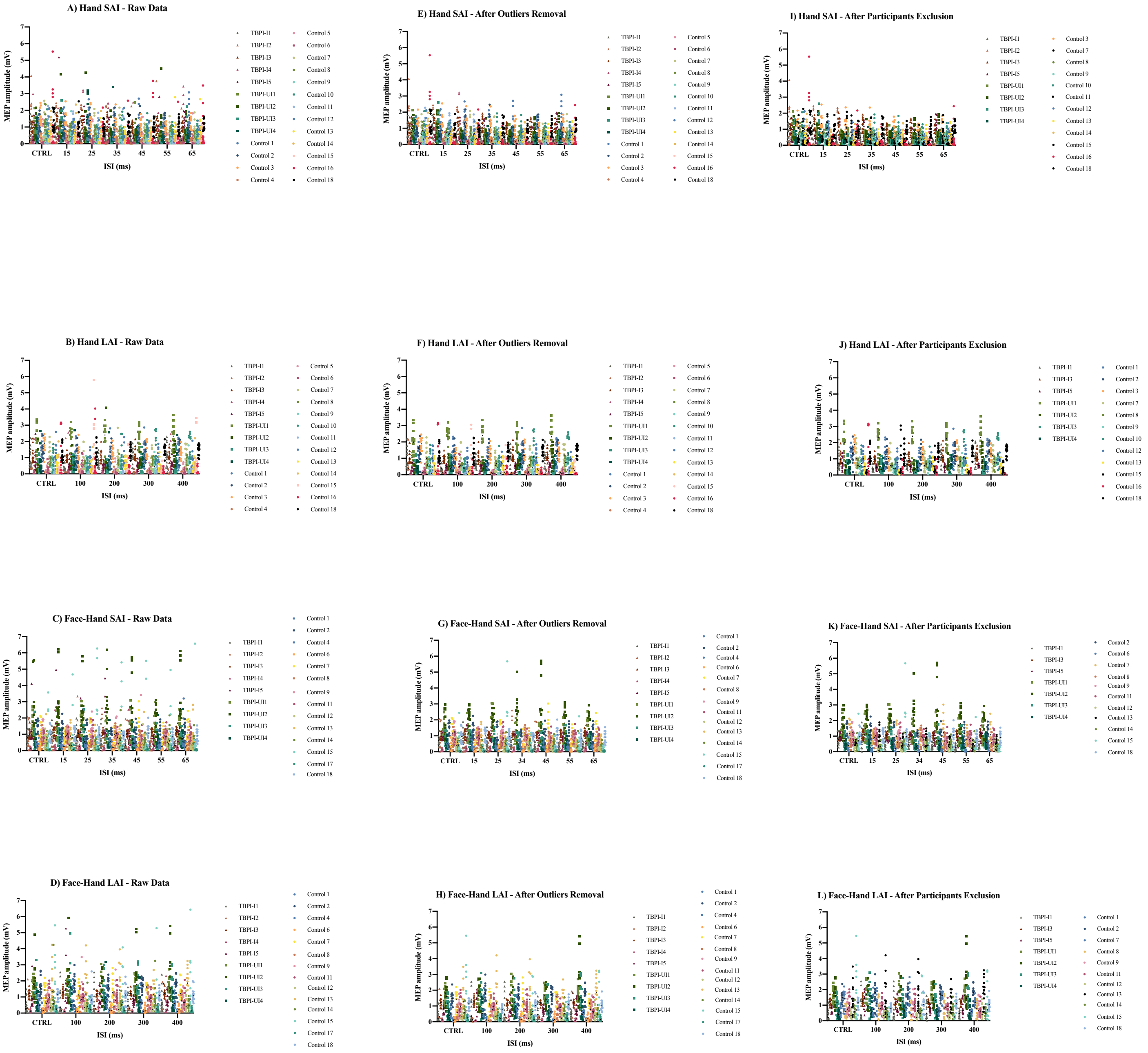

### Supplemental Figure 2

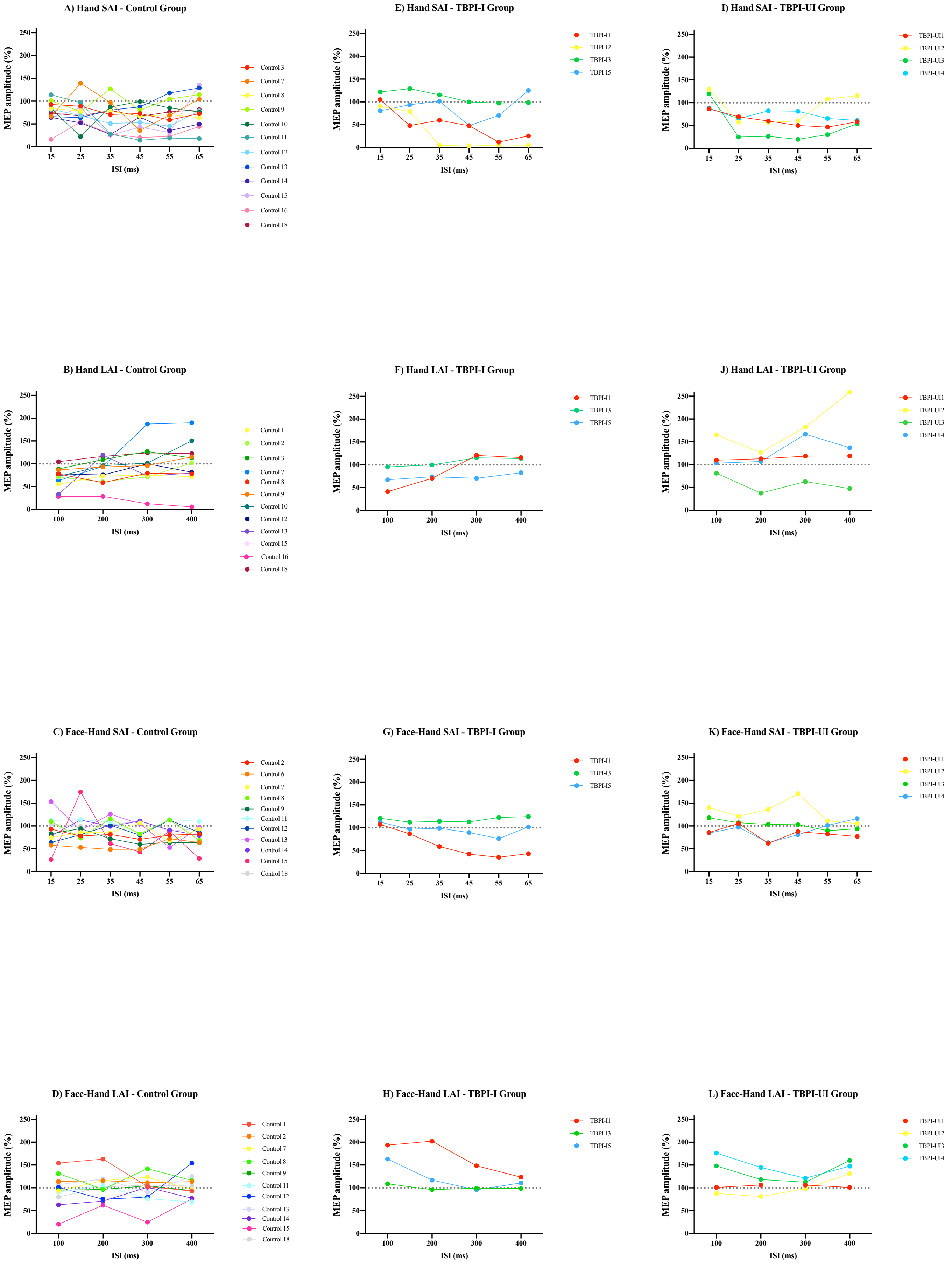
